# Using parametric g-computation to estimate the effect of long-term exposure to air pollution on mortality risk and simulate the benefits of hypothetical policies: the Canadian Community Health Survey cohort (2005 to 2015)

**DOI:** 10.1101/2023.02.06.23285546

**Authors:** Chen Chen, Hong Chen, Aaron van Donkelaar, Richard T. Burnett, Randall V. Martin, Li Chen, Michael Tjepkema, Megan Kirby-McGregor, Yi Li, Jay S. Kaufman, Tarik Benmarhnia

## Abstract

**Background:** Numerous epidemiological studies have documented the adverse health impact of long-term exposure to fine particulate matter (PM_2.5_) on mortality even at relatively low levels. However, methodological challenges remain to consider potential regulatory intervention’s complexity and provide actionable evidence on the predicted benefits of interventions. We propose the parametric g-computation as an alternative analytical approach to such challenges.

**Method:** We applied the parametric g-computation to estimate the cumulative risks of non-accidental death under different hypothetical intervention strategies targeting long-term exposure to PM_2.5_ in the Canadian Community Health Survey cohort from 2005 to 2015. On both relative and absolute scales, we explored benefits of hypothetical intervention strategies compared to the natural course that 1) set the simulated exposure value at each follow-up year to a threshold value if exposure was above the threshold (8.8 µg/m^3^, 7.04 µg/m^3^, 5 µg/m^3^, and 4 µg/m^3^); and 2) reduce the simulated exposure value by a percentage (5% and 10%) at each follow-up year. We used the three-year average PM_2.5_ concentration with one-year lag at the postal code of respondents’ annual mailing addresses as their long-term exposure to PM_2.5_. We considered baseline and time-varying confounders including demographics, behavior characteristics, income level, and neighborhood socioeconomic status. We also included the R syntax for reproducibility and replication.

**Results:** All hypothetical intervention strategies explored led to lower 11-year cumulative mortality risks than the estimated value under natural course without intervention, with the smallest reduction of 0.20 per 1000 participants (95% CI: 0.06 to 0.34) under the threshold of 8.8 µg/m^3^, and the largest reduction of 3.40 per 1000 participants (95% CI: -0.23 to 7.03) under the relative reduction of 10% per interval. The reductions in cumulative risk, or numbers of deaths that would have been prevented if the intervention was employed instead of maintaining status quo, increased over time but flattened towards the end of follow-up. Estimates among those ≥65 years were greater with a similar pattern. Our estimates were robust to different model specifications.

**Discussion:** We found evidence that any intervention further reducing the long-term exposure to PM_2.5_ would reduce the cumulative mortality risk, with greater benefits in the older population, even in a population already exposed to low levels of ambient PM_2.5_. The parametric g-computation used in this study provides flexibilities in simulating real world interventions, accommodates time-varying exposure and confounders, and estimates adjusted survival curves with clearer interpretation and more information than a single hazard ratio, making it a valuable analytical alternative in air pollution epidemiological research.

## Introduction

As collective efforts in previous decades have successfully reduced the level of fine particulate matter (PM_2.5_) globally, quantifying the effectiveness of policies that further reduce ambient PM_2.5_ is becoming particularly important in supporting evidence-based policymaking. Indeed, previous studies found consistent evidence of deleterious associations between long-term exposure to low levels of PM_2.5_ (e.g., below the current health-based standards or guidelines) and risk of mortality^1–6^ and morbidity,^7–9^ suggesting potential reductions in health burden if the PM_2.5_ level were to be further reduced. While the evaluation of exposure-response functions in existing studies provides important information in understanding the potential effectiveness of policies, further methodological considerations are required to estimate the potential benefits of realistic interventions.

First, evidence suggested that the risk associated with the changes in acute exposure to PM_2.5_ could vary with time,^10–13^ potentially due to changes in chemical compositions of PM_2.5_ with different toxicity and population susceptibility towards PM_2.5_.^14,15^ Similar disparity in toxicity across long-term exposure to PM_2.5_ components was also observed,^16,17^ suggesting that such temporal changes could exist in risk associated with long-term exposure to PM_2.5_. In other words, it is important to use analytical methods flexible enough to incorporate such temporal changes in estimation of related health burdens. However, existing studies of health impacts of long-term exposure to PM_2.5_ generally considered time-fixed exposure and confounders (see Table S1 for a narrative review of recent studies on health impact of long-term exposure to PM_2.5_ and their methodological considerations). Furthermore, the most widely used estimate for exposure-response function in this field is a single hazard ratio (HR) for the follow-up period estimated with a standard Cox-proportional hazard model (Table S1), which is assumed to be constant over time and precludes consideration of temporal changes. Although extension of a Cox-proportional hazard model could provide period-specific HRs that incorporate temporal changes,^18^ recent developments in causal inference literature raise concern about the ambiguity in the causal interpretation of HR and period-specific HRs.^19^ Specifically, period-specific HRs have a built-in selection bias because susceptible people exposed to higher PM_2.5_ are more likely to die early if PM_2.5_ truly increases risk of mortality, and are removed from the susceptible population in later time.^20^ This differential depletion of susceptible subjects over time can lead to artificially diminished or even reversed period-specific HR later in study even when the cumulative survival is still lower among those exposed to higher PM_2.5_, violating the proportional assumption and hindering interpretation.^21^

Second, calculation of health burden related to long-term exposure to PM_2.5_ commonly employed exposure-response function previously estimated with the static intervention strategy, where a fixed change of exposure value was assigned to the entire population.^22(chap19)^ However, the more flexible and realistic dynamic intervention strategy, where the exposure value was assigned based on individuals’ history of covariates including exposure, is hard to apply when existing exposure-response functions are used.^22(chap19)^ Methods capable of incorporating dynamic intervention strategy to imitate complexities in actual regulatory interventions are needed to provide direct evidence on effectiveness of air pollution control policies.^23^ To fill this gap in knowledge translation, we propose the parametric g-computation as an analytical alternative in air pollution epidemiological research, which could better predict the effectiveness of hypothetical policies while being more flexible in resembling real world interventions.

G-computation (also known as g-formula) is a generalization of non-parametric standardization developed under the potential outcome framework for causal inference,^24^ and parametric g-computation is a variation that employs parametric modeling. Under the consistency (i.e., the exposure is defined unambiguously, and all exposed individuals receive the same version of treatment),^22(chap3),25^ exchangeability (i.e., no unmeasured confounding or informative censoring),^25^ and positivity (i.e., probability of receiving every exposure conditioning on confounders is greater than zero) assumptions,^22(chap3)^ and a time-to-event outcome setting, g-computation can provide marginal causal risk estimates at each follow-up time point under hypothetical intervention strategies (i.e. adjusted survival curves), while allowing other population characteristics to be altered according to the intervention.^26^ Particularly, parametric g-computation excels in estimating adjusted survival curves under dynamic intervention strategies. In other words, g-computation can directly answer causal questions such as: how many lives could we save if we promulgate a policy that further reduces air pollution to levels lower than the current standard among those whose exposure were above the current standard, compared to maintaining the status quo? Although parametric g-computation has been widely applied in other fields of epidemiology,^27–30^ application in air pollution studies remains limited. Previous applications in this field either focused on a small cohort in occupational settings,^31–33^ or modelled simple air pollution changes on asthmatic outcomes among children (i.e. not considering time-varying confounding nor changes in effect estimates over time).^34,35^

In this study, we aim to demonstrate the use of parametric g-computation to evaluate the effectiveness of hypothetical intervention strategies targeting long-term exposure to PM_2.5_ on reducing mortality using a Canadian cohort experiencing low PM_2.5_ exposure from 2005 to 2015. This analytical alternative can account for previously unaddressed complexities, refine the effect estimates with less restrictive identification conditions and provide estimates more intuitive to policy makers.

## Methods

### Study population

We created a retrospective cohort with respondents to the Canadian Community Health Survey (CCHS) from three enrolling cycles in the years of 2000/2001, 2003 and 2005, respectively.^36–38^ CCHS is a national cross-sectional survey collecting health status, heath care utilization and health determinants information of the Canadian population, covering the population 12 years and over in the ten provinces and the three territories. The survey excluded individuals living on reserves and other Aboriginal settlements, full-time members of the Canadian Forces, the institutionalized population, and residents of certain remote regions.

Among CCHS respondents who gave permission to share and link their information with other administrative datasets, we obtained their mailing address history and death records through December 31^st^, 2015 via Statistics Canada’s Social Data Linkage Environment, using probabilistic methods based on common identifiers.^2,39^ We focused on non-accidental death as outcome (International Classification of Diseases ninth revision codes 001 to 799 and International Classification of Diseases tenth revision codes A to R) in this study. To facilitate pooling of results across cycles, we restricted the cohort to participants who were alive on January 1^st^, 2005 and used this date as the start of follow-up for all cycles. We also restricted our cohort to individuals older than 25 years and younger than 80 years in 2005 thus all cohort participants were adults and followed for 11 years or till death. Besides, we dropped respondents without data for covariates including exposure in 2005. This study was approved by the Health Canada-Public Health Agency of Canada Research Ethics Board.

### Exposure assessment

To estimate respondents’ long-term exposure to PM_2.5_, we utilized the ground-level PM_2.5_ concentrations from V4.NA.02.MAPLE of the Atmospheric Composition Analysis Group of Washington University,^40^ which covers all of North America below 70°N. The 0.01° × 0.01° (roughly equivalent to 1×1km^2^ at the latitudes where most Canadians live) annual estimates of PM_2.5_ from 2001 to 2015 were derived using satellite retrievals of aerosol optical depth and chemical transport model simulations, and calibrated with ground-based observations using geographically weighted regression.^41^ The annual estimates of PM_2.5_ closely agree with long-term cross-validated ground measurements at fixed-site monitors (n=2,312) across North America (R^2^=0.70).^41^ Using the ground-level PM_2.5_ concentration surfaces described above, we first assigned the annual PM_2.5_ concentration of the grid cell into which the postal code centroid falls as the postal code specific annual PM_2.5_ concentrations. Then we calculated respondents’ annual long-term exposure to PM_2.5_ as three-year average postal code specific PM_2.5_ concentrations with one-year lag based on their mailing address history (e.g., a respondent’s long-term exposure to PM_2.5_ in 2013 is the average of their postal code specific PM_2.5_ concentrations in 2010, 2011 and 2012).^2^ We utilized three-year average with one-year lag to represent long-term exposure of PM_2.5_ so that the exposure always precedes the outcome and the timeframe is consistent with the regulatory review of Canadian Ambient Air Quality Standards for annual PM_2.5_.^42^ This metric of long-term exposure to PM_2.5_ was widely used in previous Canada based studies of long-term health impacts of PM_2.5_.^2,43,44^

### Covariates other than exposure

In this section we summarized the data sources and meaning of covariates in this study while the covariate selection to control for in our model will be discussed in the statistical analysis section. We used covariates to describe the collection of exposure, time-fixed confounders, and time-varying confounders in this study. Baseline characteristics of respondents were ascertained at the time of enrollment into CCHS via self-report and were processed using the same method as previous studies,^2,43^ including sex, age (converted to value in 2005), body mass index, marital status, immigrant status, visible minority, indigenous status, smoking status, alcohol consumption, consumption of fruits and vegetables, leisure physical activity, working status, and educational attainment (details of variable categorization in Table 1). By using 2005 as the start of follow-up time for all individuals, we assumed that all baseline characteristics other than age ascertained at the time of enrollment would remain the same through the entire follow-up period.

**Table 1.** Descriptive statistics for participants of the Canadian Community Health Survey cohort at the start of follow-up (2005) by cycle.

| Characteristics | Cycle 2000/2001 | Cycle 2003 | Cycle 2005 |
| --- | --- | --- | --- |
| Cohort size | 62365<sup>a</sup> | 62380<sup>a</sup> | 66835<sup>a</sup> |
| Non-accidental deaths (%) | 6475 (10.4) | 6525 (10.5) | 6135 (9.2) |
| **Time-fixed covariates** |  |  |  |
| Age [mean (SD)], years | 52.1 (13.4) | 52.1 (14.4) | 50.9 (14.9) |
| Sex, % |  |  |  |
| Female | 45.2 | 45.9 | 46.2 |
| Male | 54.8 | 54.1 | 53.8 |
| BMI, % |  |  |  |
| Normal weight (18.5–24.9) | 37.5 | 32.2 | 32.0 |
| Overweight (25.0–29.9) | 36.7 | 39.8 | 39.8 |
| Obese 1 (30.0–34.9) | 16.4 | 19.1 | 18.9 |
| Obese 2 ($\geq 35$) | 6.8 | 8.1 | 8.4 |
| Underweight ($< 18.5$) | 2.6 | 0.8 | 0.9 |
| Marital status, % |  |  |  |
| Married or common-law | 65.9 | 64.3 | 63.0 |
| Separated, widowed, or divorced | 19.6 | 20.8 | 20.8 |
| Single | 14.5 | 14.9 | 16.2 |
| Immigrant status, % |  |  |  |
| Immigrant | 10.7 | 11.3 | 11.6 |
| Time lived in Canada among immigrants [mean (SD)], years | 37.4 (13.3) | 36.8 (13.9) | 35.7 (14.1) |
| Non-immigrant | 89.3 | 88.7 | 88.4 |
| Visible minority status, % |  |  |  |
| Visible minority | 5.4 | 6.3 | 4.4 |
| Not a visible minority | 94.6 | 93.7 | 95.6 |
| Indigenous status, % |  |  |  |
| Indigenous | 1.8 | 2.3 | 0<sup>b</sup> |
| Non-indigenous | 98.2 | 97.7 | 100 |
| Smoking status, % |  |  |  |
| Never smoker | 26.8 | 27.6 | 29.0 |
| Occasional smoker | 44.5 | 47.7 | 47.1 |
| Smoke under 10 cigarettes/day | 3.8 | 4.3 | 4.2 |
| Smoke 11–20 cigarettes/day | 6.0 | 5.6 | 5.7 |
| Smoke 20+ cigarettes/day | 10.9 | 9.0 | 8.6 |
| Former smoker | 8.0 | 5.8 | 5.4 |
| Alcohol consumption, % |  |  |  |
| Never drinker | 4.4 | 4.2 | 4.1 |
| Occasional drinker | 13.1 | 13.7 | 13.6 |
| Regular drinker, binge unknown | 20.3 | 18.7 | 18.2 |
| Regular, non-binge drinker | 29.4 | 31.0 | 30.4 |
| Regular, binge drinker | 26.9 | 26.7 | 27.3 |
| Former drinker | 5.9 | 5.7 | 6.4 |
| Daily consumption of fruits and vegetables, % |  |  |  |
| Under 5 servings/day | 64.7 | 59.6 | 29.9 |
| 5–10 servings/day | 32.4 | 37.0 | 19.6 |
| 10+ servings/day | 2.9 | 3.4 | 1.6 |
| Choose to not answer<sup>c</sup> | NA | NA | 48.9 |
| Employment status, % |  |  |  |
| Employed | 66.6 | 62.3 | 61.8 |
| Not employed | 2.6 | 2.6 | 2.3 |
| Not in work force | 30.8 | 35.1 | 35.9 |
| Education, % |  |  |  |
| No high school diploma | 24.1 | 22.4 | 20.6 |
| High school | 18.9 | 18.0 | 15.4 |
| Any post-secondary | 42.0 | 42.5 | 46.1 |
| University | 15.0 | 17.1 | 17.9 |
| Leisure time physical activity, % |  |  |  |
| Active | 21.2 | 24.0 | 23.4 |
| Moderately active | 25.4 | 26.4 | 26.7 |
| Inactive | 53.4 | 49.6 | 49.9 |
| Urban form, % |  |  |  |
| Active urban core | 6.2 | 7.0 | 7.0 |
| Transit reliant suburb | 3.9 | 4.3 | 4.6 |
| Auto reliant suburb or no data | 26.5 | 29.4 | 29.3 |
| Exurb | 4.8 | 5.0 | 4.8 |
| Non-CMA/CA<sup>d</sup> | 58.6 | 54.3 | 54.3 |
| Airshed, % |  |  |  |
| Western | 12.0 | 10.7 | 10.4 |
| Prairie | 16.0 | 14.9 | 13.5 |
| Western Central | 9.1 | 8.6 | 7.8 |
| Southern Atlantic | 17.2 | 14.6 | 17.3 |
| East Central | 44.1 | 49.3 | 48.9 |
| Northern | 1.6 | 1.9 | 2.1 |
| **Time-varying covariates** |  |  |  |
| Community size, % |  |  |  |
| Population > 1,500,000 | 13.5 | 14.7 | 16.9 |
| Population 500,000-1,499,999 | 10.3 | 11.9 | 10.4 |
| Population 100,000-499,999 | 20.4 | 21.0 | 19.7 |
| Population 30,000-99,999 | 14.7 | 13.2 | 12.4 |
| Population 10,000-29,999 | 7.4 | 7.0 | 7.5 |
| Non-CMA/CA<sup>d</sup> | 33.7 | 32.2 | 33.1 |
| Annual family income quintile, % |  |  |  |
| 1<sup>st</sup> quintile (lowest) | 19.0 | 19.1 | 19.3 |
| 2<sup>nd</sup> quintile | 19.7 | 19.3 | 19.1 |
| 3<sup>rd</sup> quintile | 19.9 | 19.8 | 19.5 |
| 4<sup>th</sup> quintile | 20.7 | 20.4 | 20.8 |
| 5<sup>th</sup> quintile (highest) | 20.7 | 21.4 | 21.3 |
| Canadian marginalization index—age and labour force, % |  |  |  |
| 1<sup>st</sup> quintile (lowest marginalization) | 14.4 | 15.3 | 14.8 |
| 2<sup>nd</sup> quintile | 13.5 | 13.6 | 13.7 |
| 3<sup>rd</sup> quintile | 13.7 | 13.9 | 14.1 |
| 4<sup>th</sup> quintile | 22.2 | 20.9 | 20.8 |
| 5<sup>th</sup> quintile (highest marginalization) | 36.2 | 36.3 | 36.6 |
| Canadian marginalization index—material resources, % |  |  |  |
| 1<sup>st</sup> quintile (lowest marginalization) | 15.5 | 15.7 | 15.2 |
| 2<sup>nd</sup> quintile | 16.8 | 16.9 | 17.0 |
| 3<sup>rd</sup> quintile | 20.8 | 20.8 | 20.2 |
| 4<sup>th</sup> quintile | 18.1 | 17.8 | 17.0 |
| 5<sup>th</sup> quintile (highest marginalization) | 28.8 | 28.8 | 30.6 |
| Canadian marginalization index—immigration and visible minority, % |  |  |  |
| 1<sup>st</sup> quintile (lowest marginalization) | 42.6 | 41.4 | 41.8 |
| 2<sup>nd</sup> quintile | 26.9 | 26.9 | 26.5 |
| 3<sup>rd</sup> quintile | 17.0 | 17.3 | 15.9 |
| 4<sup>th</sup> quintile | 8.5 | 9.0 | 9.9 |
| 5<sup>th</sup> quintile (highest marginalization) | 5.0 | 5.4 | 5.9 |
| Canadian marginalization index—households and dwellings, % |  |  |  |
| 1<sup>st</sup> quintile (lowest marginalization) | 22.7 | 21.2 | 21.7 |
| 2<sup>nd</sup> quintile | 28.3 | 27.7 | 27.1 |
| 3<sup>rd</sup> quintile | 21.0 | 21.7 | 21.0 |
| 4<sup>th</sup> quintile | 17.3 | 17.8 | 17.6 |
| 5<sup>th</sup> quintile (highest marginalization) | 10.7 | 11.6 | 11.8 |
| Average PM<sub>2.5</sub> of previous three years [mean (SD)], µg/m<sup>3</sup> | 6.4 (2.2) | 6.5 (2.3) | 6.5 (2.3) |
| Average PM<sub>2.5</sub> of previous three years [median (minimum, 25<sup>th</sup> percentile, 75<sup>th</sup> percentile, maximum)], µg/m<sup>3</sup> | 5.9 (1.7, 15.0, 4.6, 7.8) | 6.1 (1.7, 4.7, 8.1, 15.0) | 6.1 (1.6, 4.6, 8.2, 15.0) |

Note: BMI, Body Mass Index; PM<sub>2.5</sub>, particulate matter < 2.5 µm diameter.
<sup>a</sup>Rounded to the nearest 5 or 0 in the last digit to protect privacy
<sup>b</sup>We did not include the indigenous status indicator in models of Cycle 2005.
<sup>c</sup>Consumption of fruit and vegetable was listed with an additional option in Cycle 2005 but not in the other two cycles.
<sup>d</sup>Not categorized as census metropolitan area (CMA) or census agglomeration status (CA) and likely in rural area.

We also obtained characteristics of the respondents and their neighborhoods through linkage with administrative datasets using similar methods as previous studies.^2,43^ Specifically, we obtained annual income quintile of respondents through linkage with tax data based on common identifiers.^43^ For person-years with missing annual family income, we imputed them with the nearest prior values and the proportions of missing are 5.21%, 4.97% and 4.69% for Cycle 2000/2001, 2003 and 2005, respectively. We also obtained annual characteristics of neighborhoods through linkage with census data from the nearest census year based on respondents’ mailing address postal codes, including community size at census metropolitan area level and four Canadian Marginalization Index at census dissemination area level. The Canadian Marginalization Index summarizes dissemination area-level socioeconomic status into four dimensions using principal component analysis to reduce the dimensionality of census data: the immigration and visible minority index combines information on proportion of recent immigrants and proportion of people self-identify as visible minority; the households and dwellings index combines information on types and density of residential accommodations and family structure characteristics; the material resources index combines information on access to and attainment of basic material needs; and the age and labour force index combines information on participation in labour force and proportion of seniors.^45^ Last, we obtained airshed (six distinct regions of Canada that cut cross jurisdictional boundaries and showed similar air quality characteristics and air movement patterns within each region) to capture large scale spatial variation,^46^ and urban form information of respondents’ neighborhoods in 2005 to capture urbanicity of participants’ residence, through linkage with census data.^2^

### Hypothetical intervention strategies

In this study, we explored three types of intervention strategies: 1) applying the simulated value of time-varying covariates without any intervention (natural course); 2) setting the simulated long-term exposure to PM_2.5_ value at each follow-up year to a threshold value if PM_2.5_ was higher than the threshold (threshold intervention); and 3) reducing the simulated PM_2.5_ value by a fixed percentage at each interval (i.e., follow-up year) (relative reduction intervention).

Threshold values explored are the current Canadian Ambient Air Quality Standards for PM_2.5_ of 8.8 µg/m^3^, 80% of the current Canadian Ambient Air Quality Standards for PM_2.5_ (or 7.04 µg/m^3^), the new World Health Organization (WHO) air quality guideline of 5 μg/m^3^, and a PM_2.5_ level that was further below the WHO guideline (4 µg/m^3^). Interval-specific relative reduction values explored are 10% and 5% per interval. To avoid extensive model extrapolation, we restricted the relative reduction intervention so that subjects with exposure below 1.8 µg/m^3^, the background PM_2.5_ level in Canada,^47^ will not be further reduced. The first type of intervention strategy represents the predicted covariates based on the observed data without intervening and serves as the reference for other strategies. The second and the third are dynamic intervention strategies that incorporate the impact of intervention on covariates during earlier time points while simulating covariates in later time points.

### Statistical analysis

We applied parametric g-computation with different hypothetical intervention strategies targeting long-term exposure to PM_2.5_ to understand the benefits of intervention strategies on cumulative risk of non-accidental death. We conducted g-computation analysis for each enrollment cycle separately and pooled the results across cycles using meta-regression. Briefly, we estimated the cumulative mortality risk at each follow-up year standardized to the distribution of the confounders and long-term exposure to PM_2.5_ in the study population, with all time-varying covariates (confounders and PM_2.5_) conditioned on covariates history, with and without intervention on PM_2.5_ (i.e., adjusted survival curves). Next, we calculated the differences in cumulative mortality risks between the natural course and other intervention strategies on both absolute and relative scales to provide estimates for the benefits of hypothetical intervention strategies compared to maintaining status quo. We pooled results with fixed-effect meta-regression, which calculates a weighted average of cycle specific estimates with weights equal to the inverse of the variance using the “meta” package.^48^

The proof of parametric g-computation are described extensively elsewhere,^22(chap21),29^ and detailed description of how to implement such an approach in a setting similar to our study was previously published,^28^ with available R package for easy implementation.^49^ However, since the application of parametric g-computation is limited in air pollution studies, we include a diagram (Figure 1) to summarize the four steps that carry out the g-computation in a time-to-event setting with time-varying exposure and confounders, and describe the steps in details below.

**Figure 1.**
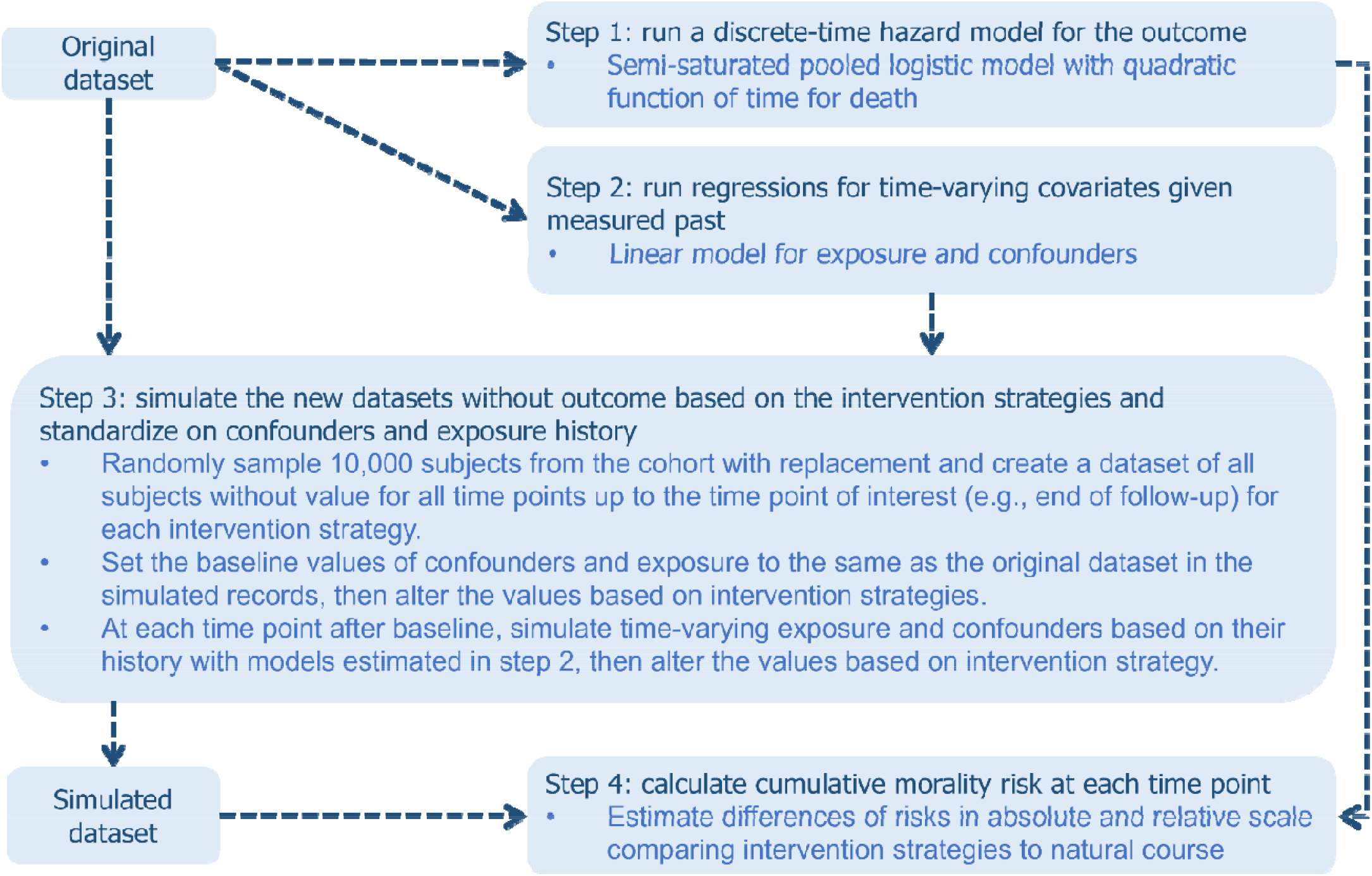
Diagram of g-computation with time-to-event outcome and time-varying covariates. Arrow indicates information needed from previous box.

### Steps to implement parametric g-computation

In Step 1, we fitted a pooled logistic model (i.e., discrete-time hazard model) and adjusted for baseline characteristics, time-varying characteristics, quadratic function of year and interaction between long-term exposure to PM_2.5_ and categorical year. The pooled logistic model estimated the probability of death during the year conditioning on survival till the start of the year given all covariates (including PM_2.5_), which allowed the conditional probability of death and its association with PM_2.5_ to vary over year. We chose confounders to control for in the outcome model based on substantive knowledge of the relationship between long-term PM_2.5_ and mortality as summarized in the simplified directed acyclic graph (Figure S1). We included a full list of covariates in Table 1 with specific forms of covariates in Table 2. We included both individual socioeconomic status indicators (e.g., education and family income) and community socioeconomic status indicators (e.g., Canadian Marginalization Index for dissemination area) to fully capture the variation in socioeconomic status among cohort participants, which is a major source of residual confounding. We also included individual behavior indicators like dietary and exercise patterns, which are strong risk factors for mortality, precede the exposure, and might share common unmeasured causes with the exposure, even though they might not directly cause the exposure.^50^ To note, in the setting when only time-fixed covariates were used, we could estimate marginal adjusted survival curves directly using outputs from this pooled logistic model by predicting the probability of death standardized to the distributions of covariates under the intervention of interest (e.g., setting the baseline level of exposure to a specific value while keeping all baseline covariates the same as observed for all participants).^19,51^ However, in our study setting of time-varying covariates and time-to-event outcome, we also need to model time-varying covariates (including PM_2.5_) so that we could simulate time-varying covariates at all follow-up years for all participants, especially for periods after participants’ death.^28,29^

**Table 2.** Details for covariate formats and model types for both outcome and covariate models in main analysis.

| Variable Name | Type as independent variable | Type as dependent variable and corresponding model used |
| --- | --- | --- |
| **Time-fixed covariates** |  |  |
| Age in 2005<sup>a</sup> | Restricted cubic spline function with 5 knots | Not predicted |
| Sex | Binary | Not predicted |
| BMI | 5 categories | Not predicted |
| Marital status | 3 categories | Not predicted |
| Immigrant | An indicator for immigrant and a interaction term between the indicator and a continuous variable for years in Canada among immigrants | Not predicted |
| Visible minority | Binary | Not predicted |
| Indigenous status | Binary | Not predicted |
| Smoking status | 6 categories | Not predicted |
| Alcohol consumption | 6 categories | Not predicted |
| Daily consumption of fruit and vegetables | 4 categories | Not predicted |
| Leisure time physical activity | 3 categories | Not predicted |
| Employment status | 3 categories | Not predicted |
| Education attainment | 4 categories | Not predicted |
| Urban form | 5 categories | Not predicted |
| Air shed | 6 categories | Not predicted |
| **Time-varying covariates** |  |  |
| Time | Year and quadratic term of Year<sup>b</sup> | Not predicted |
| Community size | Continuous | Bounded normal<sup>c</sup> (1 to 6) and linear regression |
| Annual family income quintile | Continuous | Bounded normal (1 to 5) and linear regression |
| Canadian Marginalization Index for immigration and visible minority | Continuous | Bounded normal (1 to 5) and linear regression |
| Canadian Marginalization Index for material resources | Continuous | Bounded normal (1 to 5) and linear regression |
| Canadian Marginalization Index for households and dwellings | Continuous | Bounded normal (1 to 5) and linear regression |
| Canadian Marginalization Index for age and labour force | Continuous | Bounded normal (1 to 5) and linear regression |
| Three-year average PM<sub>2.5</sub> concentration with one-year lag | Natural logarithm transformed | Normal with linear regression |

Note: BMI, Body Mass Index; PM<sub>2.5</sub>, particulate matter < 2.5 µm diameter.
<sup>a</sup>In subset analysis restricted to cohort participants older or equal to 65 years, we used restricted cubic spline function with 3 knots for age.
<sup>b</sup>Categorical year was used in the interaction terms between time and the exposure.
<sup>c</sup>Variables with bounded normal category was modeled and simulated by using the standardized value (subtracting the minimum value and dividing by the range) in linear regression. Simulated values that fall outside the observed range are set to the minimum or maximum of the observed range.

In Step 2, we modeled the time-varying covariates (including PM_2.5_) using linear regressions while including variables such as previous-year value of the covariate of interest, baseline characteristics, same-year values of time-varying covariates set to occur before the covariate of interest, and quadratic function of time. The choice of independent variables in covariate models are based on substantive knowledge as summarized in the simplified directed acyclic graph (Figure S1). We summarized the list of all covariates in Table 1 and the specific forms of covariates included in covariate models in Table 2. We set the sequence of time-varying covariates as community size, income, immigration and visible minority, material resources, households and dwellings, age and labour force and long-term exposure to PM_2.5_. Since previous studies using different cycles of CCHS found a supra-linear PM_2.5_-mortality association,^2,43,52,53^ we used natural logarithm transformed long-term exposure to PM_2.5_ as the independent variable in both the outcome and covariate model in the main analysis.

In Step 3, we simulated new datasets based on the intervention strategies. For each intervention, we randomly sampled 10,000 subjects from the cohort with replacement (i.e., Monte Carlo sampling) and created an empty dataset of all sampled subjects for all follow-up years till the end of period of interest (normally the end of follow-up as in this study but extrapolation is possible with extra assumptions). We only simulated new datasets for 10,000 subjects instead of the number of participants in study cohorts (~60,000 participants in each cohort) to save computation time and similar practice was conducted before with smaller cohort.^29^ Next, we assigned the baseline values of all covariates (values of baseline covariates and values of time-varying covariates at start of follow-up) in each simulated dataset to the same as its original dataset, then altered the relevant covariate values based on the intervention strategy (e.g., setting the baseline long-term exposure to PM_2.5_ to 5 μg/m^3^ if it is higher than 5 μg/m^3^ in the threshold intervention of 5 μg/m^3^ but could include other covariates if needed). Last, we simulated time-varying covariates at each year after baseline based on their history with covariate models estimated in the second step and altered the covariates based on the intervention strategy.

In Step 4, with the simulated datasets and outcome model from the first step, we calculated for each subject the probability of dying during each year conditioning on surviving to the beginning of this year, standardized to the distribution of the confounders and long-term exposure to PM_2.5_ under the intervention strategies, regardless of their observed outcomes. Next, we calculated for each subject the cumulative mortality risk at each year as the cumulative sum of the abovementioned conditional probability of mortality times the probability of surviving till the beginning of the time interval. The estimated cumulative morality risk at each year is the average of estimates from all subjects for all hypothetical interventions. We also calculated the absolute difference in cumulative morality risk and percentage change in cumulative morality risk with estimated cumulative morality risk from natural course as reference.

Besides, we calculated the 95% confidence intervals (CI) for all estimates using standard errors from 200 bootstrap iterations.^54^ In each iteration, we randomly sampled the same number of participants as in the original cohort with replacement and ran the four steps described above to calculate cumulative mortality risks under intervention strategies. We chose this number of iteration because we were constrained by available computational resources (>1h of computational time for each bootstrap iteration), and the original application of parametric g-computation in time-varying covariates and time-to-event setting used the same number.^29^ Future studies with more computational resources might consider larger number of bootstrap iteration.

### Sensitivity analyses

To test the robustness of our results to model misspecification, we considered a number of different model specifications for both outcome and covariate models including: 1) reordering the sequence of time-varying covariates in covariate models by moving age and labour force to before the other Canadian Marginalization Index, moving income to after Canadian Marginalization Index, and moving PM_2.5_ to the first place among all covariates; 2) extending the extent of history modeled by including previous year and two-year previous values of all the time-varying covariates in the covariate models other than just the previous-year value of the covariate of interest; and 3) including time-varying covariates other than long-term PM_2.5_ as categorical in the outcome model and using the multinomial logistic model for them in covariate model instead of modeling them as continuous with bounds using linear model (see Table 2 for details of model specifications for each time-varying covariates in main analysis). We also visually evaluated the simulated and observed adjusted survival curves and histories of covariates under no intervention in the main analysis as a heuristic check of model misspecification.^27^

Next, to facilitate comparison with previous studies, we used long-term PM_2.5_ in original scale in all models as sensitivity analysis, which assumed the same log-linear PM_2.5_-mortality association used in other cohorts^4,7^ instead of the supra-linear one supported by previous studies of different cycles of the CCHS cohort.^2,43,52,53^ Besides, we also ran a Cox-proportional hazard model with the same specification as the outcome model in our main analysis except that we included no time variable and used long-term PM_2.5_ in original scale, which assumed a log-linear PM_2.5_-mortality association.

Last, since most deaths occurred among older individuals and age could modify the health impact of long-term exposure to PM_2.5_, we conducted a subset analysis restricted to cohort participants aged ≥65 years at the time of enrollment. Since it took up to 24 hours to run one round of sensitivity analysis without bootstrapping, we were unable to perform bootstrapping to calculate CIs for sensitivity analyses due to computational constraints and did not present variances for our estimates. We pooled cycle-specific estimates from sensitivity analyses by averaging the estimates in each cycle. All analyses were done in R version 4.0.5^55^ with the “gfoRmula” package.^49^ The R code to replicate these analyses and a simulated dataset are available at the following link: https://github.com/suthlam/cchs_g_computation.git.

## Results

We observed 6,475 (10.4%), 6,525 (10.5%), and 6,135 (9.2%) non-accidental deaths in the 11 years of follow-up starting from 2005 among the three cycles of CCHS cohorts of 62,365, 62,380, and 66,385 participants, respectively (Table 1). The three cycle cohorts were comparable in all descriptive statistics (Table 1). Without any hypothetical intervention, the observed average long-term exposure to PM_2.5_ in three cycles of our cohort decreased slightly from 6.4 ± 2.2 µg/m^3^, 6.5 ± 2.3 µg/m^3^, 6.5 ± 2.3 µg/m^3^ in 2005 to 5.8 ± 2.0 µg/m^3^, 6.0 ± 2.0 µg/m^3^, and 6.0 ± 2.0 µg/m^3^ in 2015, respectively (Table 1).

All hypothetical intervention strategies explored in this study led to lower 11-year cumulative mortality risks than the estimated value under a natural course without intervention, 102.5 per 1000 participants (95% confidence interval (CI): 100.3 to 104.8 per 1000 participants). The reductions in 11-year cumulative mortality risks from the natural course were 0.20 per 1000 participants (95% CI: 0.06 to 0.34 per 1000 participants) under the threshold of 8.8 µg/m^3^, 0.63 per 1000 participants (95% CI: 0.18 to 1.07 per 1000 participants) under the threshold of 7.04 µg/m^3^, 1.87 per 1000 participants (95% CI: 0.53 to 3.21 per 1000 participants) under the threshold of 5 µg/m^3^, 3.08 per 1000 participants (95% CI: 0.85 to 5.31 per 1000 participants) under the threshold of 4 µg/m^3^, 1.68 per 1000 participants (95% CI: -0.15 to 3.51 per 1000 participants) under the relative reduction of 5% per interval, and 3.40 per 1000 participants (95% CI: -0.23 to 7.03 per 1000 participants) under the relative reduction of 10% per interval. To note, the reduction in 11-year cumulative mortality risks could also be interpreted as the number of deaths that would have been prevented if the intervention was employed instead of maintaining status quo. Changes in relative scale showed similar pattern (Table 3). To fulfill the four threshold intervention strategies, averages of 18.7%, 38.3%, 72.0% and 91.4% of subjects experienced change in their natural course exposure every year, respectively, while 100% had their exposure changed under the two relative reduction intervention strategies (Table 3). The corresponding reductions in average simulated PM_2.5_ from the start of follow-up to the end of year 11 ranged from 0.13 to 1.87 µg/m^3^ for threshold intervention strategies, and 1.25 to 2.18 µg/m^3^ for relative reduction intervention strategies (Table 3). Across all strategies, we observed steady expansions in differences of yearly cumulative mortality risks between the natural course and other strategies until the 7^th^ year of follow-up, after which the differences remain constant and shrink during the last year of follow-up (Figure 2 with numeric results in Table S2). In the main analysis, we pooled estimates of yearly cumulative mortality risks across cycles using random-effect meta-regression and pooled estimates of differences (absolute and relative scale) in cumulative mortality risks using fixed-effect meta-regression. Cycle-specific results with corresponding I^2^ values are summarized in Figure S2 with numeric results in Table S3.

**Table 3.** Summaries of estimated 11-year cumulative mortality risk under different intervention strategies pooled across cycles and differences in estimated risk compared to natural course in relative and absolute scale, and corresponding average simulated PM_2.5_ and proportion of subjects with exposure changed for all intervention strategies.

| **Intervention strategy** | **11-year CMR ( per 1000 participants) (95% CI)** | **Difference in 11-year CMR ( per 1000 participants) (95% CI)** | **Percentage change in 11-year CMR (95% CI)** | **Average percentage of subjects with exposure changed<sup>a</sup>** | **Average simulated PM<sub>2.5</sub> concentration (µg/m<sup>3</sup>)<sup>a</sup>** |
| --- | --- | --- | --- | --- | --- |
| Natural course | 102.5 (100.3, 104.8) | Reference | Reference | 0 | 5.62 |
| Threshold of 8.8 µg/m<sup>3</sup> | 102.3 (100.1, 104.6) | -0.20 (-0.34, -0.06) | -0.19 (-0.33, -0.05) | 18.7 | 5.49 |
| Threshold of 7.04 µg/m<sup>3</sup> | 102.0 (99.7, 104.2) | -0.63 (-1.07, -0.18) | -0.60 (-1.03, -0.17) | 38.3 | 5.21 |
| Threshold of 5 µg/m<sup>3</sup> | 100.9 (98.4, 103.5) | -1.87 (-3.21, -0.53) | -1.79 (-3.11, -0.48) | 72.0 | 4.42 |
| Threshold of 4 µg/m<sup>3</sup> | 99.8 (96.7, 102.9) | -3.08 (-5.31, -0.85) | -2.95 (-5.14, -0.77) | 91.4 | 3.75 |
| 5% reduction per interval | 101.4 (98.6, 104.2) | -1.68 (-3.51, 0.15) | -1.61 (-3.40, 0.17) | 100 | 4.37 |
| 10% reduction per interval | 99.8 (95.6, 103.9) | -3.40 (-7.03, 0.23) | -3.27 (-6.81, 0.28) | 100 | 3.44 |

Note: CMR: Cumulative Mortality Risk; PM<sub>2.5</sub>, particulate matter < 2.5 µm diameter.
<sup>a</sup>This is the three-cycle average of the mean value across all years.

**Figure 2.**
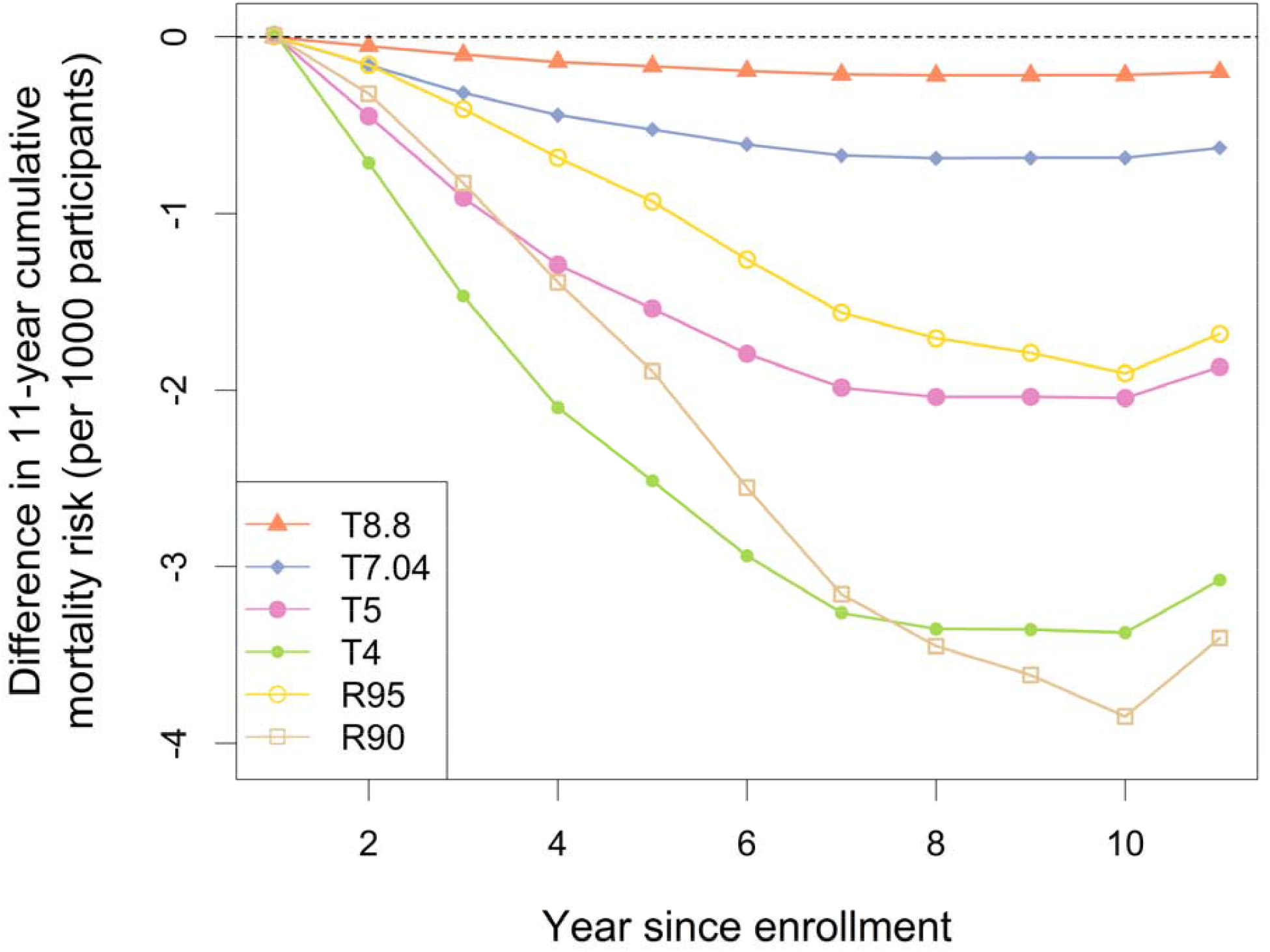
Differences in yearly cumulative mortality risks pooled across cycles comparing different intervention strategies to natural course, with weights equal to the inverse of variance. Numeric results are presented in Table S2. Note: T8.8, threshold value (reduced to threshold value if above) set at the current Canadian Ambient Air Quality Standards for PM_2.5_ of 8.8 µg/m^3^; T7.04: threshold value set at 80% of the current Canadian Ambient Air Quality Standards for PM_2.5_ (or 7.04 µg/m^3^); T5: threshold value set at the new World Health Organization guideline of 5 µg/m^3^; T4: threshold value set at a PM_2.5_ level that was further below the World Health Organization guideline (4 µg/m^3^); R90: yearly relative reduction values set at 10% per interval; and R95: yearly relative reduction values set at 5% per interval.

Heuristic checks of model fitting in the main analysis support the robustness of our estimates: 1) the cumulative mortality risk estimated by parametric g-computation under the natural course closely tracked the value observed (Figure S3); and 2) the observed mean values of all time-varying covariates were similar to those simulated under the natural course over time (Figure S3). To note, since participants had no time-varying covariates recorded after their death while we simulated participants’ time-varying covariates for all years, differences between observed and simulated covariates are expected, especially later in the study period. Furthermore, sensitivity analyses with different model specifications (different sequence of time-varying covariate, extent of history modeled, and parametrization of time-varying confounders) resulted in similar estimates as the main analysis (Figure 3, numeric results in Table S4).

**Figure 3.**
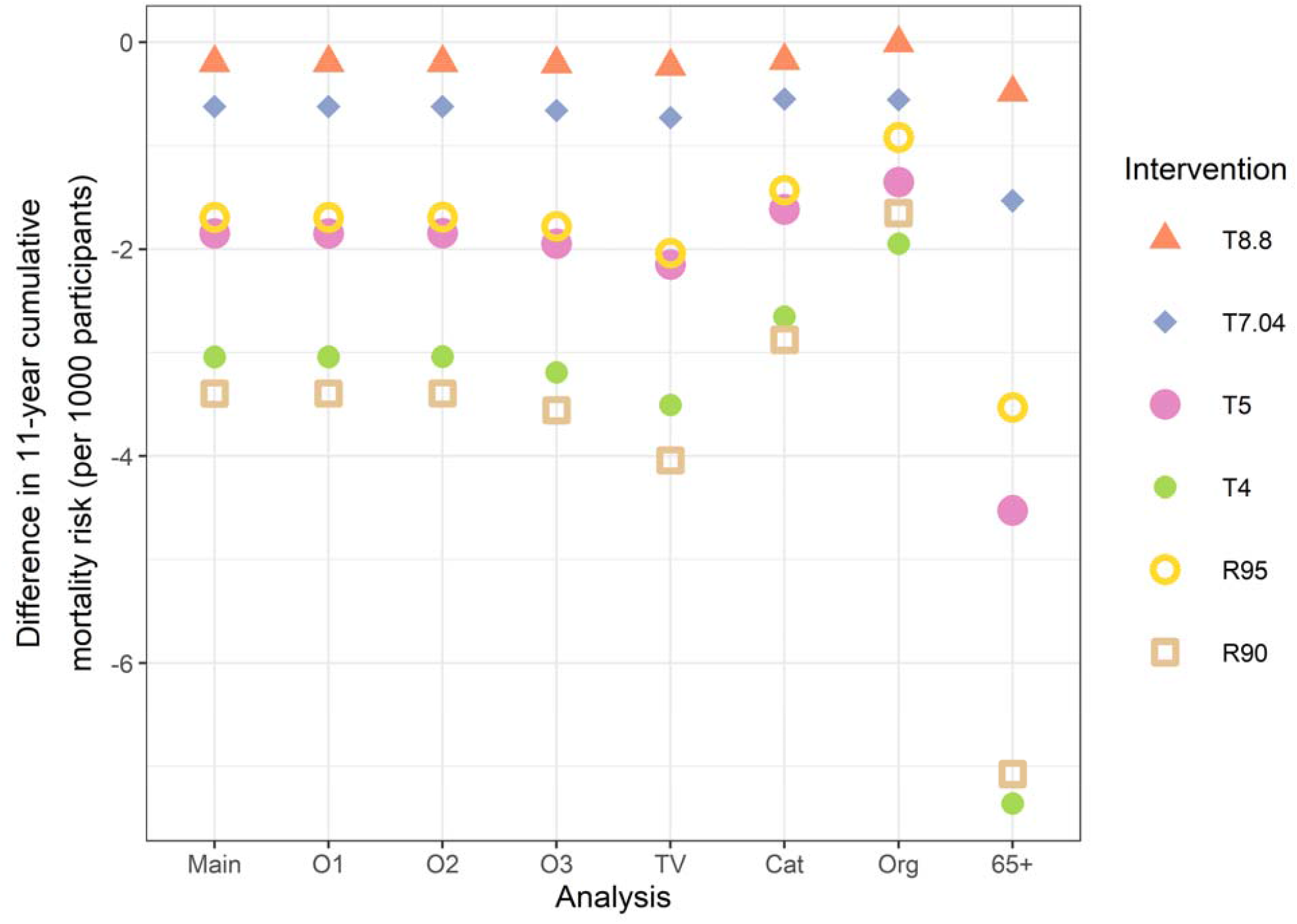
Differences in 11-year cumulative mortality risks comparing different intervention strategies to natural course for main analysis and sensitivity analyses. Numeric results are presented in Table 3 and Table S4. Note: T8.8, threshold value (reduced to threshold value if above) set at the current Canadian Ambient Air Quality Standards for PM_2.5_ of 8.8 µg/m^3^; T7.04: threshold value set at 80% of the current Canadian Ambient Air Quality Standards for PM_2.5_ (or 7.04 µg/m^3^); T5: threshold value set at the new World Health Organization guideline of 5 µg/m^3^; T4: threshold value set at a PM_2.5_ level that was further below the World Health Organization guideline (4 µg/m^3^); R90: yearly relative reduction values set at 10% per interval; R95: yearly relative reduction values set at 5% per interval; O1: placing Canadian Marginalization Index-age and labour force before the other Canadian Marginalization Index in occurring sequence of time-varying covariate; O2: moving income to after Canadian Marginalization Index in occurring sequence of time-varying covariate; O3: moving PM_2.5_ to the first in occurring sequence of time-varying covariate; TV: adding all time-varying covariates of previous year and two-year previous to covariate model; Cat: including time-varying covariates other than long-term PM_2.5_ as categorical in outcome model and using multinomial logistic model for them in covariate model; Org: using long-term PM_2.5_ in original scale in all models; and 65+: subset analysis restricted to cohort participants older or equal to 65 years.

When assuming a log-linear PM_2.5_-mortality association in the sensitivity analysis (compared to the supra-linear association assumed in main analysis by log-transforming the exposure), reductions in 11-year cumulative mortality risks comparing other intervention strategies to the natural course ranged from 0.01 per 1000 participants to 1.65 per 1000 participants, slightly smaller than main analysis assuming a supra-linear PM_2.5_-mortality association (log-transformed PM_2.5_ used as exposure in modeling) (Table S4). The Cox model assuming a log-linear PM_2.5_-mortality association found 15.6% (95% CI: 4.0 to 28.5%) increase in hazard of death per 10 µg/m^3^ increase in PM_2.5_. When focusing on cohort participants ≥65 years at the start of follow-up, reductions in 11-year cumulative mortality risks comparing other intervention strategies to the natural course ranged from 0.49 per 1000 participants to 7.07 per 1000 participants (Table S4), which is larger than the main analysis using the general population ≥35 years.

## Discussion

In this study, we applied the parametric g-computation as an analytical alternative that is particularly valuable for air pollution epidemiological research, especially for evaluating specific intervention strategies. With application in a large Canadian cohort, we demonstrated how to incorporate consideration of complex time structure in the data and how to calculate causally interpretable cumulative risk estimates over the follow-up time (i.e., adjusted survival curves) with parametric g-computation. We described that any intervention further reducing the long-term exposure to PM_2.5_ would reduce the cumulative mortality risk, even in a region with relatively low levels of ambient PM_2.5_. Such reduction in cumulative risk increased over time and flattened towards the end of follow-up on both relative and absolute scales. The older population also experienced greater benefits from the explored hypothetical intervention strategies than the general population.

Numerous observational studies found positive associations between long-term exposure to PM_2.5_ and non-accidental mortality. A meta-analysis reported a pooled effect estimate of 6% (95% CI: 4 to 8%) increase in hazard of death per 10 µg/m^3^ increase in PM_2.5_ (HR-1).^5^ A recent study in a similar Canadian cohort from 2000 to 2012 found 11% (95% CI: 4 to 18%) increase in hazard of non-accidental death per 10 µg/m^3^ increase in chronic exposure to PM_2.5_ with a Cox proportional hazard model.^2^ Our sensitivity analysis using Cox model without time-varying coefficients found similar numeric results [15.6% (95% CI: 4.0 to 28.5%)]. Although we can’t directly compare our estimates from the main analysis to previous results given the difference in interventions explored, the consistent reductions in cumulative mortality risk over follow-up time across intervention strategies when compared to natural course in this study lend further support to previous findings that PM_2.5_ is detrimental to health even at levels below current standards.

For example, we identified a 0.19% (95% CI: 0.05 to 0.33%) decrease in 11-year cumulative mortality risk comparing the hypothetical intervention strategy with threshold of 8.8 µg/m^3^ to natural course, which provided evidence of health benefits from policies that further reduce the air pollution level to below current Canada standard of 8.8 µg/m^3^, which is lower than the 12 µg/m^3^ standard of U.S. explored by previous studies.^4,9^ To facilitate comparison with previous studies assuming a log-linear PM_2.5_-mortality association, we included sensitivity analysis using PM_2.5_ on the normal scale and found reduced cumulative mortality risks in all hypothetical interventions compared to maintaining status quo but the numeric values are smaller than those in the main analysis. The observed difference in the numeric values of analysis assuming log-linear association and analysis assuming supra-linear association is a combination of difference in how the exposure-response relationship is modeled and how the exposure model performs. However, given the existing evidence in Canadian cohorts and similarity between observed survival curve and estimated survival curve using parametric g-computation under no intervention in the main analysis,^2,43,52,53^ we have confidence in the validity of results assuming a supra-linear association.

More importantly, we demonstrated how to incorporate more flexibilities in simulating real world interventions with g-computation in this study and provide intuitive estimates for benefits of such interventions. Taking the hypothetical intervention strategy with threshold of the current Canadian Ambient Air Quality Standards as an example, the average long-term exposure to PM_2.5_ in 2005 was around 6.5 µg/m^3^, below the standard of 8.8 µg/m^3^. However, some cohort participants were exposed to PM_2.5_ levels higher than 8.8 µg/m^3^ during some years of follow-up and our hypothetical intervention only affected these subject-years by reducing their exposure to 8.8 µg/m^3^, which represented a three-cycle average of 18.7% of subjects across all years. Since the observed PM_2.5_ levels decreased without any intervention in our study, fewer subjects were directly intervened on in later years under threshold intervention strategies, which explained the flattened differences in cumulative risks between intervention strategies in later years. However, all time-varying covariates after the intervention on PM_2.5_ would change accordingly due to the intervention on PM_2.5_, thus influencing future outcomes as well. Such dynamic intervention strategy incorporated considerations of people who could be intervened on and are more realistic than the static intervention strategy commonly employed in health burden estimation with traditional exposure-response function, which sets change in exposure at a fixed value for all individuals throughout the period of interest. Besides, although we only provided differences in cumulative risk as compared to the natural course, it is easy to estimate differences between any two hypothetical intervention strategies.

Furthermore, the estimated cumulative risks over the follow-up time by g-computation (i.e., adjusted survival curves) and corresponding comparisons of values between different hypothetical interventions provided clearer causal interpretation and more information than a single HR or period-specific HRs, which is generally used in air pollution studies (Table S1). In the context of health impacts from chronic exposure to PM_2.5_, HR can change over time since the toxicity of PM_2.5_ (e.g., chemical composition of PM_2.5_) and susceptibility of population to PM_2.5_ could change over time, while standard Cox model assumed constant HR and period-specific HR from extensions of Cox had built-in bias that led to ambiguity in causal interpretation.^56^ On the other hand, the cumulative mortality risks estimated in this study avoided such ambiguity in interpretation while also demonstrating the change of intervention effect over time.^19^ Also, obtaining the casually interpretable absolute differences in cumulative risks between hypothetical intervention strategies over time could be particularly helpful for comparing different scenarios regarding public health benefits.^57^ Besides, if policies affecting air pollutants such as PM_2.5_ could further affect prognostic covariates influencing both future PM_2.5_ levels and health outcomes (commonly referred to as exposure-confounder feedback), traditional regression methods based on adjustment in a multivariable model would fail and lead to biased estimates for the effect while g-computation is designed to particularly solve this problem.^24,26,58^ An example of such exposure-confounder feedback is that people might move due to high level of PM_2.5_ in their current community and subsequently change the characteristics of their community of residence, while the characteristics of their current community also affect the level of PM_2.5_ and probability of death. Controlling for such community characteristics is necessary for confounding control but doing so with traditional methods will remove the indirect effect mediated through community characteristics and introduce collider-stratification bias^59^ if any unmeasured confounder of the community characteristics and death exists.^58^ However, making moving decision based on community level of PM_2.5_ is unlikely in countries with relatively low PM_2.5_ like Canada and exposure-confounder feedback is expected to be negligible in our study, but it is possible in countries with higher PM_2.5_.

This study has a few limitations that need to be acknowledged. First, parametric g-computation can only account for measured confounders and lack of conditional exchangeability (i.e., residual confounding) might exist due to unmeasured confounders or measurement errors of existing confounders, regardless of our extensive list of confounders considered based on substantive knowledge on risk factors of PM_2.5_ exposure and death (Figure S1). For example, we assumed many individual behavior, demographic, and socioeconomic variables to be time-invariant (e.g., employment status and body mass index) due to data availability (these variables were only reported once at the time of enrollment) while they likely changed over the study period.

However, we also included time-varying individual income and community demographic and socioeconomic variables in our models, which mitigated the concern of residual confounding from these baseline variables. Besides, like other cohort studies of air pollution, we utilized postal-code level PM_2.5_ levels as surrogates for individual exposure to PM_2.5_, which might introduce exposure misclassification.^60^ Recent studies showed that such bias may either not bias effect estimates ^61^ or bias these estimates towards the null,^62^ making our estimates more conservative.

Second, although we explored different model specifications and found similar results in sensitivity analyses, we cannot rule out the possibility of model misspecification, especially given the fact that parametric g-computation requires correct model specification of both the outcome and covariate models to achieve unbiased results. Notably, McGrath et al. demonstrated that the “g-null paradox”, a form of model misspecification that was traditionally believed to cause false rejection of null hypothesis under sharp null effect,^63^ is unavoidable in parametric g-computation even when the sharp null hypothesis does not hold, and recommended more flexible models in analysis.^64^ However, the magnitude of bias depends on the extent of exposure-confounder feedback and time-varying confounding. In the context of this study, we would expect relatively small exposure-confounder feedback thus less concern over g-null paradox.

Also, consistent results from sensitivity analysis using more flexible models supported the robustness of our results.

Third, being an active research field, the existing R package for parametric g-computation does not support features like incorporation of spline functions of time-varying covariates in the model, direct estimation of randomized interventional strategy,^65^ model fit checking with significance tests, or bias analysis. However, the current package provided enough flexibility for our study to employ flexible models that mitigated the possibility of violating the positivity assumption via model extrapolation. For example, we were able to incorporate flexible supra-linear PM_2.5_-mortality association and temporal changes in the conditional probability of mortality in the estimation as supported by previous studies, incorporate restricted cubic spline function of baseline age in all models, and conduct sensitivity analysis with categorical time-varying confounders. Besides, although not relevant to our cohort since we had the all-cause mortality as the outcome and no loss to follow-up, right censoring and informative loss to follow-up could be handled by parametric g-computation and the existing R package by simulating data on participants as though they had not been censored.^66^It is worth mentioning that other methods could also handle the methodological considerations that g-computation addresses—consideration of complex time structure and reporting of adjusted survival curves— and have been applied in air pollution epidemiological research, including Inverse Probability of Treatment Weighting (IPTW).^6^ Furthermore, some recent approaches such as the targeted maximum likelihood estimation can also be used to directly evaluate individualized dynamic intervention strategies of continuous exposures and provide doubly robust estimates that are less vulnerable to model misspecification with valid statistical inference when data-adaptive/machine-learning methods are incorporated.^67,68^

Finally, PM_2.5_ is a mixture of varying chemical components and toxicity and is generated from different sources of emissions (e.g. traffic, industries, and wildfires). In this paper, we focused on PM_2.5_ without distinguishing the PM_2.5_ composition nor the sources of emissions. This potentially violated the consistency assumption (i.e. no-multiple-versions-of-treatment and all exposed individuals received the same version of treatment). If there is any unmeasured confounder of the “version of treatment” and outcome relationship, the effect estimates could be biased according to a recent simulation study, with magnitude and direction of such bias depending on the strength of confounding.^69^ In future studies, it would be important to consider the possible differential toxicity of PM_2.5_ components and define hypothetical interventions targeting different sources of PM_2.5_ emissions separately.

## Conclusion

This study demonstrated the benefits of using parametric g-computation as an analytical alternative for air pollution epidemiological research, especially for evaluating the potential effects of realistic dynamic intervention strategies in the time-to-event setting with time-varying exposure and confounders. With a large Canadian cohort, we calculated causally interpretable cumulative risk estimates over the follow-up time and corresponding benefits compared to maintaining status quo. We also found that any intervention further reducing the long-term exposure to PM_2.5_ would reduce the cumulative mortality risk from maintaining the status quo, even in a population already exposed to relatively low levels of ambient PM_2.5_.

## Data Availability

The original data could not be shared to protect the privacy of study participants. All data produced in the present work are contained in the manuscript.

