## Supplementary materials for "Using parametric g-computation to estimate the effect of long-term exposure to air pollution on mortality risk and simulate the benefits of hypothetical policies: the Canadian Community Health Survey cohort (2005 to 2015)"

Table S1. Illustrative list of studies using cohort design to explore health impacts of chronic exposure to PM_2.5_. Summary is based on the main analysis unless otherwise specified. Note: proportional hazard (PH); hazard ratio (HR).

| **Study** | **Analytical method** | **Follow-up period** | **Effect measure** | **Survival curves** | **Time-varying estimates (effect modification by time)** | **Considered complex time structure** | | | **Simulation for intervention** |
| --- | --- | --- | --- | --- | --- | --- | --- | --- | --- |
|  |  |  |  |  |  | **Time-varying exposure** | **Time-varying confounder** | **Exposure-confounder feedback** |  |
| Kioumourtzoglou et al. 2016^1^ | Cox PH model | 1999-2010 | HR | No | No | Yes | Yes | No | No |
| Di et al. 2017^2^ | Cox PH model | 2000-2012 | HR | No | No | Yes | Yes | No | No |
| Makar et al. 2017^3^ | Cox PH model with IPW | 1 year after recruitment | HR | No | No | No | No | No | No |
| Parker et al. 2018^4^ | Discrete-time PH model | 1997-2009 | HR | No | No | No | No | No | No |
| Lim et al. 2018^5^ | Cox PH model | 1995-2011 | HR | No | No | Yes, in sensitivity analysis | No | No | No |
| Garcia et al. 2019^6^ | Poisson regression | 1993-2014 | Incidence rate ratio | No | No | Yes, in sensitivity analysis | No | No | No |
| Garcia et al. 2019^7^ | Parametric g-computation (Poisson model for outcome) | 1993-2014 | Incidence rate difference | No | No | Yes, in sensitivity analysis | No | No | Yes |
| Huang et al. 2019^8^ | Cox PH model | 2000-2015 | HR | No | No | Yes | No | No | No |
| Paoin et al. 2020^9^ | Cox PH model | 2005-2013 | HR | No | No | Yes | Yes | No | No |
| Wu et al. 2020^10^ | Cox PH, Poisson regression, generalized propensity score matching / weighting / adjustment | 2000-2016 | HR | No | No | Yes | Yes | Yes | No |
| Yazdi et al. 2021^11^ | Doubly robust additive hazard model | 2000-2016 | Hazard difference | No | No | Yes | Yes | Yes | No |

Table S2. Summaries of differences in yearly cumulative mortality risks pooled across cycles comparing different intervention strategies^1^ to natural course in main analysis, with weights equal to the inverse of variance (numeric results for Figure 2).

| **Follow-up year** | **Difference in yearly cumulative mortality risk (per 1000 participants) (95% CI)** | | | | | |
| --- | --- | --- | --- | --- | --- | --- |
|  | **T8.8** | **T7.04** | **T5** | **T4** | **R95** | **R90** |
| 1 | 0.00 (0.00, 0.00) | 0.00 (0.00, 0.00) | 0.01 (-0.01, 0.03) | 0.03 (-0.01, 0.06) | 0.00 (0.00, 0.01) | 0.01 (0.00, 0.02) |
| 2 | -0.05 (-0.09, -0.02) | -0.16 (-0.27, -0.05) | -0.45 (-0.74, -0.15) | -0.71 (-1.17, -0.25) | -0.16 (-0.25, -0.06) | -0.32 (-0.51, -0.13) |
| 3 | -0.10 (-0.16, -0.05) | -0.32 (-0.49, -0.15) | -0.91 (-1.37, -0.44) | -1.47 (-2.20, -0.74) | -0.41 (-0.60, -0.22) | -0.83 (-1.2, -0.45) |
| 4 | -0.14 (-0.21, -0.07) | -0.44 (-0.65, -0.23) | -1.29 (-1.87, -0.71) | -2.1 (-3.02, -1.18) | -0.68 (-0.97, -0.40) | -1.39 (-1.96, -0.82) |
| 5 | -0.17 (-0.24, -0.09) | -0.52 (-0.77, -0.28) | -1.54 (-2.22, -0.85) | -2.51 (-3.60, -1.43) | -0.93 (-1.33, -0.53) | -1.89 (-2.70, -1.09) |
| 6 | -0.19 (-0.28, -0.10) | -0.61 (-0.89, -0.33) | -1.79 (-2.58, -1.01) | -2.94 (-4.20, -1.68) | -1.26 (-1.82, -0.70) | -2.55 (-3.67, -1.43) |
| 7 | -0.21 (-0.31, -0.11) | -0.67 (-0.98, -0.36) | -1.99 (-2.89, -1.08) | -3.26 (-4.72, -1.81) | -1.56 (-2.33, -0.79) | -3.16 (-4.68, -1.63) |
| 8 | -0.22 (-0.33, -0.11) | -0.69 (-1.03, -0.34) | -2.04 (-3.05, -1.03) | -3.35 (-4.99, -1.72) | -1.71 (-2.69, -0.73) | -3.45 (-5.40, -1.50) |
| 9 | -0.22 (-0.34, -0.10) | -0.68 (-1.06, -0.31) | -2.04 (-3.14, -0.93) | -3.36 (-5.16, -1.55) | -1.79 (-2.99, -0.59) | -3.61 (-6.00, -1.22) |
| 10 | -0.22 (-0.35, -0.09) | -0.68 (-1.09, -0.28) | -2.04 (-3.26, -0.83) | -3.37 (-5.37, -1.38) | -1.91 (-3.37, -0.44) | -3.85 (-6.76, -0.94) |
| 11 | -0.20 (-0.34, -0.06) | -0.63 (-1.07, -0.18) | -1.87 (-3.21, -0.53) | -3.08 (-5.31, -0.85) | -1.68 (-3.51, 0.15) | -3.40 (-7.03, 0.23) |

^1^Abbreviations for intervention strategies: threshold value (reduced to threshold value if above) set at the current Canadian Ambient Air Quality Standards for PM_2.5_ of 8.8 µg/m^3^ (T8.8), 80% of the current Canadian Ambient Air Quality Standards for PM_2.5_ (or 7.04 µg/m^3^) (T7.04), the new World Health Organization guideline of 5 µg/m^3^ (T5), a PM_2.5_ level that was further below the World Health Organization guideline (4 µg/m^3^) (T4), and interval-specific relative reduction values set at 10% per follow-up year (R90) and 5% per follow-up year (R95).

Table S3. Summaries of estimated 11-year cumulative mortality under different intervention strategies^1^ and differences in estimated risk compared to natural course in relative and absolute scale by cycle in main analysis.

| **Intervention strategy** | **Estimates (95% CI)** | | | **I^2^ (%)** |
| --- | --- | --- | --- | --- |
|  | **Cycle 2001/2002** | **Cycle 2003** | **Cycle 2005** |  |
|  | **11-year cumulative mortality risk (per 1000 participants)** | | |  |
| **Natural course** | 105.3 (101.2, 109.3) | 107.1 (103.1, 111.0) | 96.4 (92.7, 100) | 88.7 |
| **T8.8** | 105.0 (101, 109.1) | 107.0 (103.0, 110.9) | 96.1 (92.5, 99.8) | 88.9 |
| **T7.04** | 104.5 (100.5, 108.6) | 106.7 (102.8, 110.7) | 95.6 (91.9, 99.3) | 89.0 |
| **T5** | 103.0 (98.5, 107.4) | 106.1 (101.6, 110.5) | 94.2 (89.9, 98.5) | 87.0 |
| **T4** | 101.4 (96.1, 106.7) | 105.4 (100.1, 110.8) | 92.8 (87.5, 98.0) | 82.5 |
| **R95** | 102.8 (97.9, 107.7) | 106.5 (101.7, 111.2) | 94.4 (89.5, 99.4) | 83.6 |
| **R90** | 100.3 (93.2, 107.4) | 105.8 (98.8, 112.9) | 92.5 (85.0, 99.9) | 69.5 |
|  | **Difference in 11-year cumulative mortality risk (per 1000 participants)** | | |  |
| **T8.8** | -0.20 (-0.50, 0.00) | -0.11 (-0.36, 0.14) | -0.25 (-0.50, -0.01) | 0 |
| **T7.04** | -0.74 (-1.49, 0.01) | -0.34 (-1.13, 0.45) | -0.78 (-1.55, -0.02) | 0 |
| **T5** | -2.31 (-4.63, 0.02) | -1.01 (-3.41, 1.38) | -2.22 (-4.48, 0.04) | 0 |
| **T4** | -3.86 (-7.74, 0.03) | -1.65 (-5.62, 2.33) | -3.62 (-7.35, 0.12) | 0 |
| **R95** | -2.50 (-5.68, 0.67) | -0.61 (-3.74, 2.53) | -1.96 (-5.15, 1.22) | 0 |
| **R90** | -5.01 (-11.26, 1.24) | -1.24 (-7.54, 5.06) | -3.94 (-10.25, 2.37) | 0 |
|  | **Percentage change in 11-year cumulative mortality risk** | | |  |
| **T8.8** | -0.22 (-0.44, 0.00) | -0.10 (-0.33, 0.13) | -0.26 (-0.53, 0.00) | 0 |
| **T7.04** | -0.70 (-1.41, 0.01) | -0.32 (-1.05, 0.42) | -0.81 (-1.63, 0.01) | 0 |
| **T5** | -2.19 (-4.38, 0.00) | -0.95 (-3.18, 1.28) | -2.30 (-4.73, 0.12) | 0 |
| **T4** | -3.66 (-7.33, 0.00) | -1.54 (-5.24, 2.17) | -3.75 (-7.76, 0.26) | 0 |
| **R95** | -2.38 (-5.38, 0.62) | -0.57 (-3.50, 2.36) | -2.04 (-5.45, 1.38) | 0 |
| **R90** | -4.76 (-10.65, 1.14) | -1.16 (-7.04, 4.73) | -4.09 (-10.86, 2.68) | 0 |

^1^Abbreviations for intervention strategies: threshold value (reduced to threshold value if above) set at the current Canadian Ambient Air Quality Standards for PM_2.5_ of 8.8 µg/m^3^ (T8.8), 80% of the current Canadian Ambient Air Quality Standards for PM_2.5_ (or 7.04 µg/m^3^) (T7.04), the new World Health Organization guideline of 5 µg/m^3^ (T5), a PM_2.5_ level that was further below the World Health Organization guideline (4 µg/m^3^) (T4), and interval-specific relative reduction values set at 10% per follow-up year (R90) and 5% per follow-up year (R95).

Table S4. Summaries of estimated 11-year cumulative mortality risk under different intervention strategies^1^ pooled across all cycles and differences in estimated risk compared to natural course in relative and absolute scale in sensitivity analyses.

| **Intervention strategy** | **11-year cumulative mortality risk (per 1000 participants)** | **Difference in 11-year cumulative mortality risk (per 1000 participants)** | **Percentage change in 11-year cumulative mortality risk** | **Average percentage of subject-years with exposure changed** | **Average simulated PM_2.5_ concentration in the year 11**  **(µg/m^3^)^2^** |
| --- | --- | --- | --- | --- | --- |
| **New order of the time-varying covariates: community size, income, material resources, households and dwellings, age and labour force, immigration and visible minority, and long-term exposure to PM_2.5_** | | | | | |
| Natural course | 102.9 | Reference | Reference | 0 | 5.61 |
| T8.8 | 102.7 | -0.20 | -0.19 | 18.7 | 5.49 |
| T7.04 | 102.3 | -0.62 | -0.61 | 38.3 | 5.21 |
| T5 | 101.1 | -1.85 | -1.81 | 72.0 | 4.42 |
| T4 | 99.9 | -3.04 | -2.98 | 91.3 | 3.75 |
| R95 | 101.2 | -1.69 | -1.66 | 100 | 4.37 |
| R90 | 99.5 | -3.40 | -3.33 | 100 | 3.44 |
| **New order of the time-varying covariates: community size, immigration and visible minority, material resources, households and dwellings, age and labour force, income, and long-term exposure to PM_2.5_** | | | | | |
| Natural course | 102.8 | Reference | Reference | 0 | 5.62 |
| T8.8 | 102.6 | -0.20 | -0.19 | 18.7 | 5.49 |
| T7.04 | 102.1 | -0.62 | -0.61 | 38.3 | 5.21 |
| T5 | 100.9 | -1.85 | -1.81 | 72.0 | 4.42 |
| T4 | 99.7 | -3.04 | -2.99 | 91.3 | 3.75 |
| R95 | 101.1 | -1.69 | -1.66 | 100 | 4.37 |
| R90 | 99.4 | -3.39 | -3.34 | 100 | 3.44 |
| **New order of the time-varying covariates: long-term exposure to PM_2.5_, community size, income, immigration and visible minority, material resources, households and dwellings, and age and labour force** | | | | | |
| Natural course | 103.0 | Reference | Reference | 0 | 5.61 |
| T8.8 | 102.8 | -0.21 | -0.21 | 18.7 | 5.48 |
| T7.04 | 102.3 | -0.66 | -0.65 | 38.3 | 5.18 |
| T5 | 101.0 | -1.95 | -1.91 | 71.9 | 4.39 |
| T4 | 99.8 | -3.19 | -3.13 | 91.2 | 3.72 |
| R95 | 101.2 | -1.78 | -1.74 | 100 | 4.34 |
| R90 | 99.4 | -3.56 | -3.49 | 100 | 3.40 |
| **Added all time-varying covariates of previous year and two-year previous to covariate model** | | | | | |
| Natural course | 102.9 | Reference | Reference | 0 | 5.62 |
| T8.8 | 102.7 | -0.23 | -0.23 | 19.0 | 5.48 |
| T7.04 | 102.2 | -0.73 | -0.72 | 38.8 | 5.16 |
| T5 | 101.8 | -2.15 | -2.11 | 73.1 | 4.31 |
| T4 | 99.4 | -3.51 | -3.44 | 91.9 | 3.61 |
| R95 | 100.9 | -2.04 | -2.00 | 100 | 4.19 |
| R90 | 98.9 | -4.04 | -3.97 | 100 | 3.20 |
| **Including time-varying covariates other than long-term PM_2.5_ as categorical in outcome model and using multinomial logistic model for them in covariate model** | | | | | |
| Natural course | 101.7 | Reference | Reference | 0 | 5.61 |
| T8.8 | 101.5 | -0.18 | -0.18 | 18.8 | 5.48 |
| T7.04 | 101.2 | -0.55 | -0.55 | 38.3 | 5.20 |
| T5 | 100.1 | -1.61 | -1.61 | 71.9 | 4.42 |
| T4 | 99.0 | -2.65 | -2.65 | 91.2 | 3.75 |
| R95 | 100.3 | -1.43 | -1.43 | 100 | 4.37 |
| R90 | 98.8 | -2.87 | -2.88 | 100 | 3.44 |
| **Using long-term PM_2.5_ in original scale** | | | | | |
| Natural course | 102.9 | Reference | Reference | 0 | 5.91 |
| T8.8 | 102.9 | -0.01 | -0.01 | 1.3 | 5.91 |
| T7.04 | 102.4 | -0.56 | -0.55 | 38.0 | 5.34 |
| T5 | 101.6 | -1.35 | -1.33 | 73.9 | 4.40 |
| T4 | 101.0 | -1.95 | -1.91 | 92.6 | 3.66 |
| R95 | 102.0 | -0.92 | -0.90 | 100 | 4.63 |
| R90 | 101.3 | -1.65 | -1.62 | 100 | 3.69 |
| **Restricted to >= 65 years** | | | | | |
| Natural course | 311.1 | Reference | Reference | 0 | 5.74 |
| T8.8 | 310.6 | -0.49 | -0.16 | 20.3 | 5.60 |
| T7.04 | 309.6 | -1.53 | -0.49 | 40.6 | 5.29 |
| T5 | 306.6 | -4.53 | -1.45 | 72.9 | 4.46 |
| T4 | 303.7 | -7.36 | -2.35 | 91.9 | 3.77 |
| R95 | 307.6 | -3.53 | -1.11 | 100 | 4.46 |
| R90 | 304.0 | -7.07 | -2.23 | 100 | 3.50 |

^1^Abbreviations for intervention strategies: threshold value (reduced to threshold value if above) set at the current Canadian Ambient Air Quality Standards for PM_2.5_ of 8.8 µg/m^3^ (T8.8), 80% of the current Canadian Ambient Air Quality Standards for PM_2.5_ (or 7.04 µg/m^3^) (T7.04), the new World Health Organization guideline of 5 µg/m^3^ (T5), a PM_2.5_ level that was further below the World Health Organization guideline (4 µg/m^3^) (T4), and interval-specific relative reduction values set at 10% per follow-up year (R90) and 5% per follow-up year (R95).

^2^This is the average of all simulated subjects across three cycles of the CCHS cohort.

Figure S1. A simplified directed acyclic graph for main analysis. Note: we only kept the arrow from baseline covariates to time 0 (t0) of time-varying covariates for simplicity of the figure; we only included two time points (t0 and t1) for time-varying covariates for simplicity of the figure; we categorized the potential confounders for simplicity of the figure; demographics includes age, sex, marital status, immigrant, visible minority, and indigenous status; behavior includes smoking status, alcohol consumption, leisure physical activity and daily consumption of fruits and vegetables; individual characteristics include BMI, employment status and education; regional indicators include urban form and airshed; community characteristics includes all time-varying community level characteristics (community size and Canadian Marginalization Index).

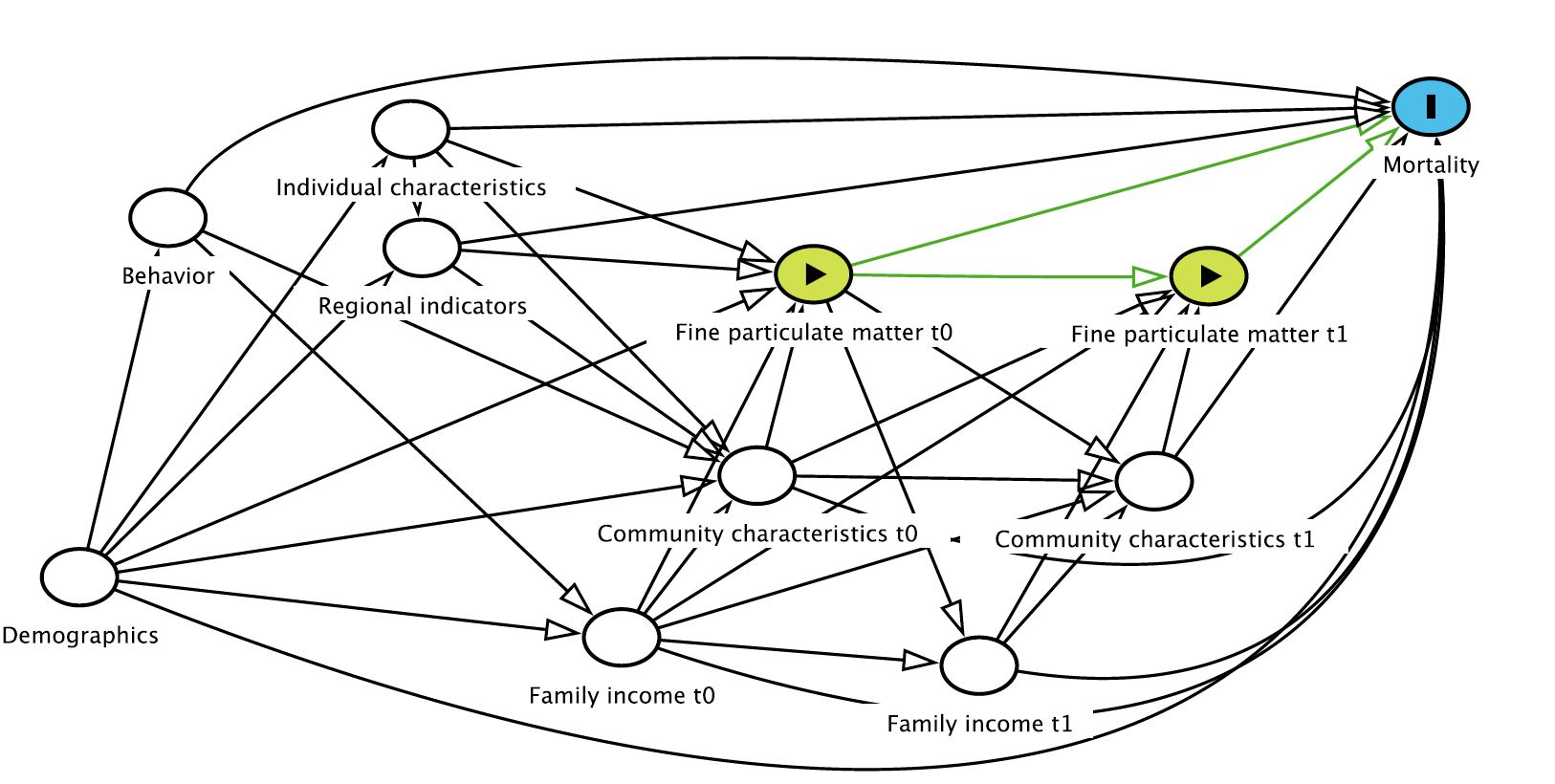

Figure S2. Estimated 11-year cumulative mortality risk under different intervention strategies and differences in estimated risk compared to natural course in relative and absolute scale by cycle. Numeric results are included in Table S3. Note: T8.8, threshold value (reduced to threshold value if above) set at the current Canadian Ambient Air Quality Standards for PM_2.5_ of 8.8 µg/m^3^; T7.04: threshold value set at 80% of the current Canadian Ambient Air Quality Standards for PM_2.5_ (or 7.04 µg/m^3^); T5: threshold value set at the new World Health Organization guideline of 5 µg/m^3^; T4: threshold value set at a PM_2.5_ level that was further below the World Health Organization guideline (4 µg/m^3^); R90: interval-specific relative reduction values set at 10% per follow-up year; and R95: interval-specific relative reduction values set at 5% per follow-up year.

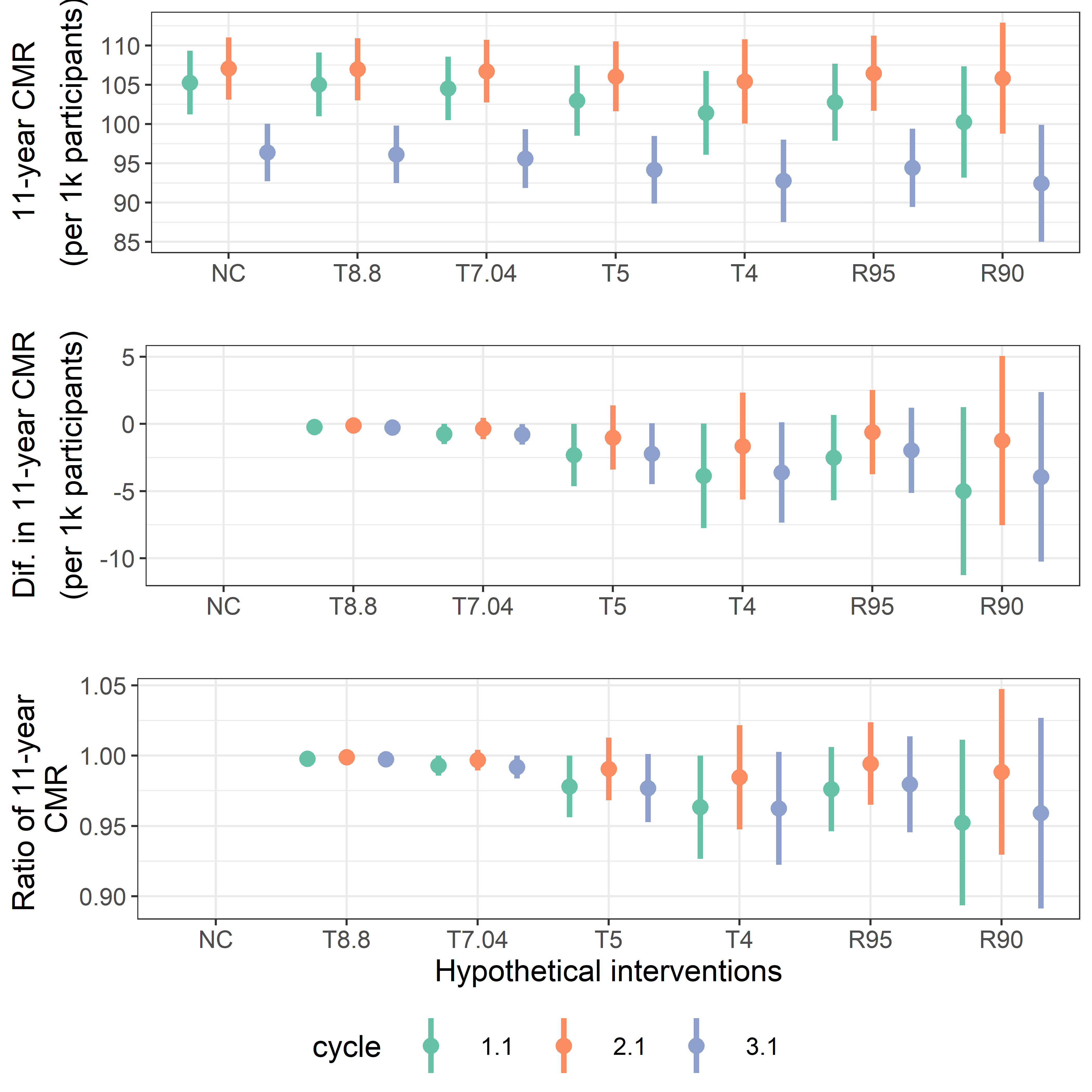

Figure S3. Comparison of observed mean vs. simulated mean under no intervention for mortality risk and time-varying covariates in each cycle. Numeric results are available upon request from the corresponding author.

**Cycle 2001/2002**

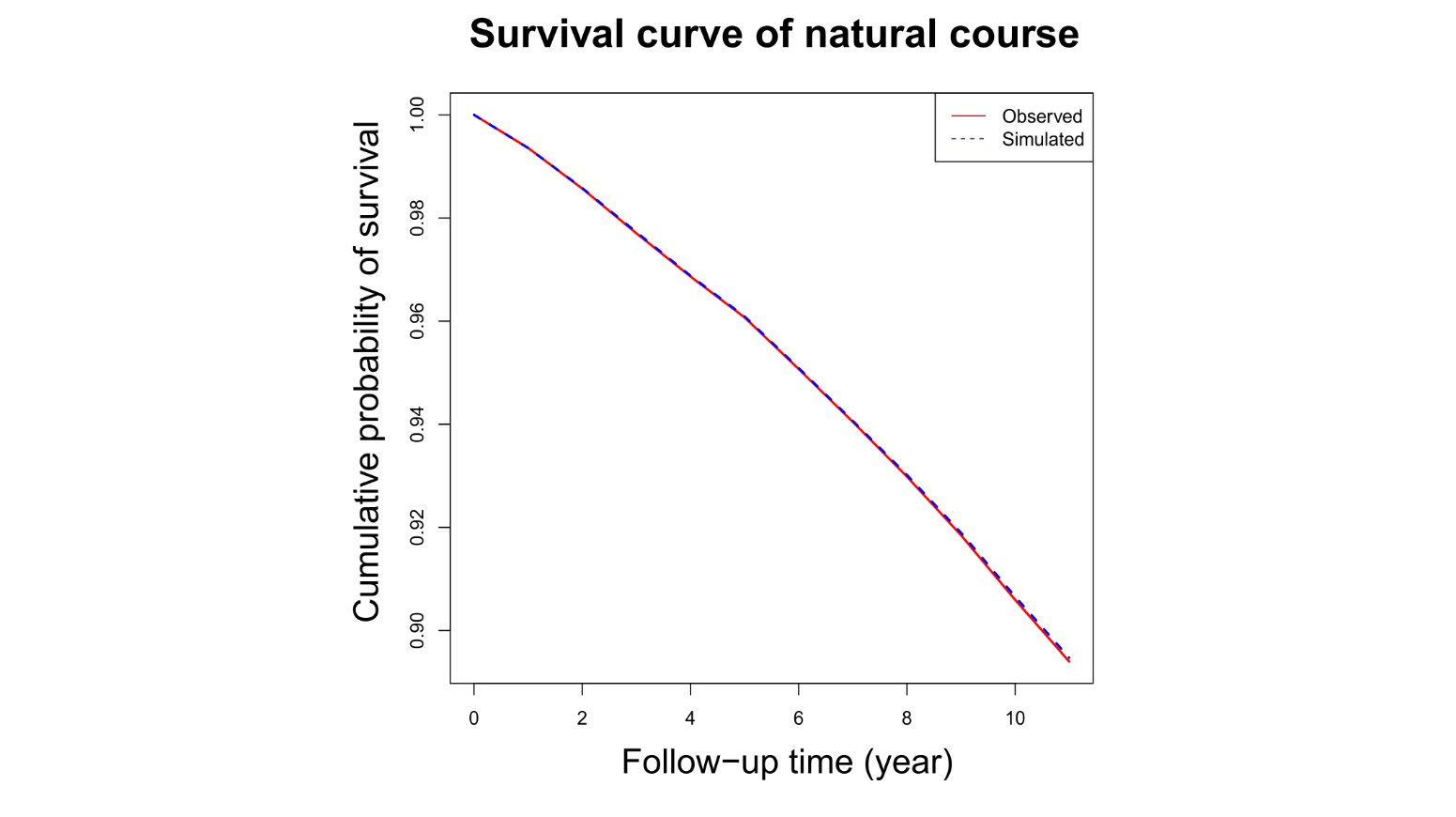

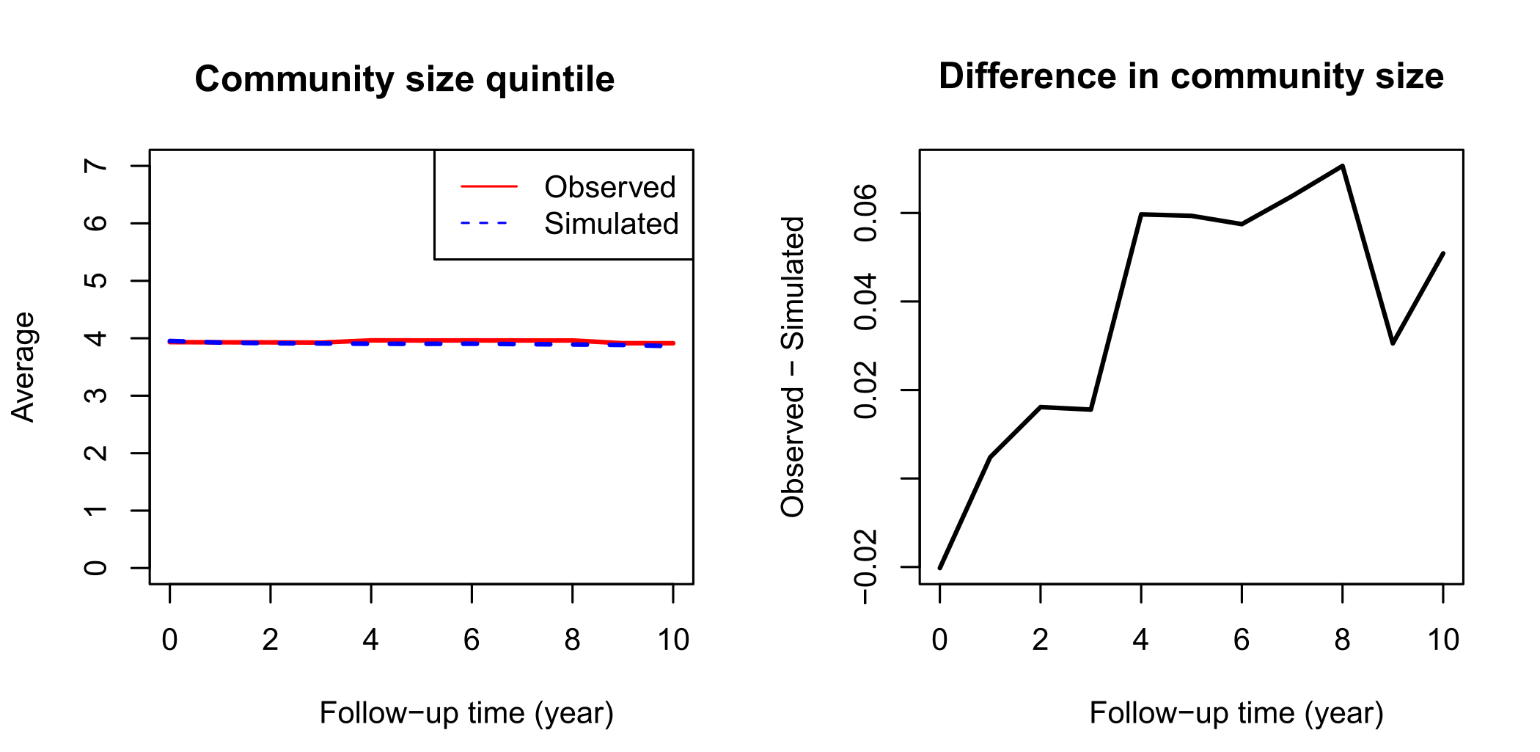

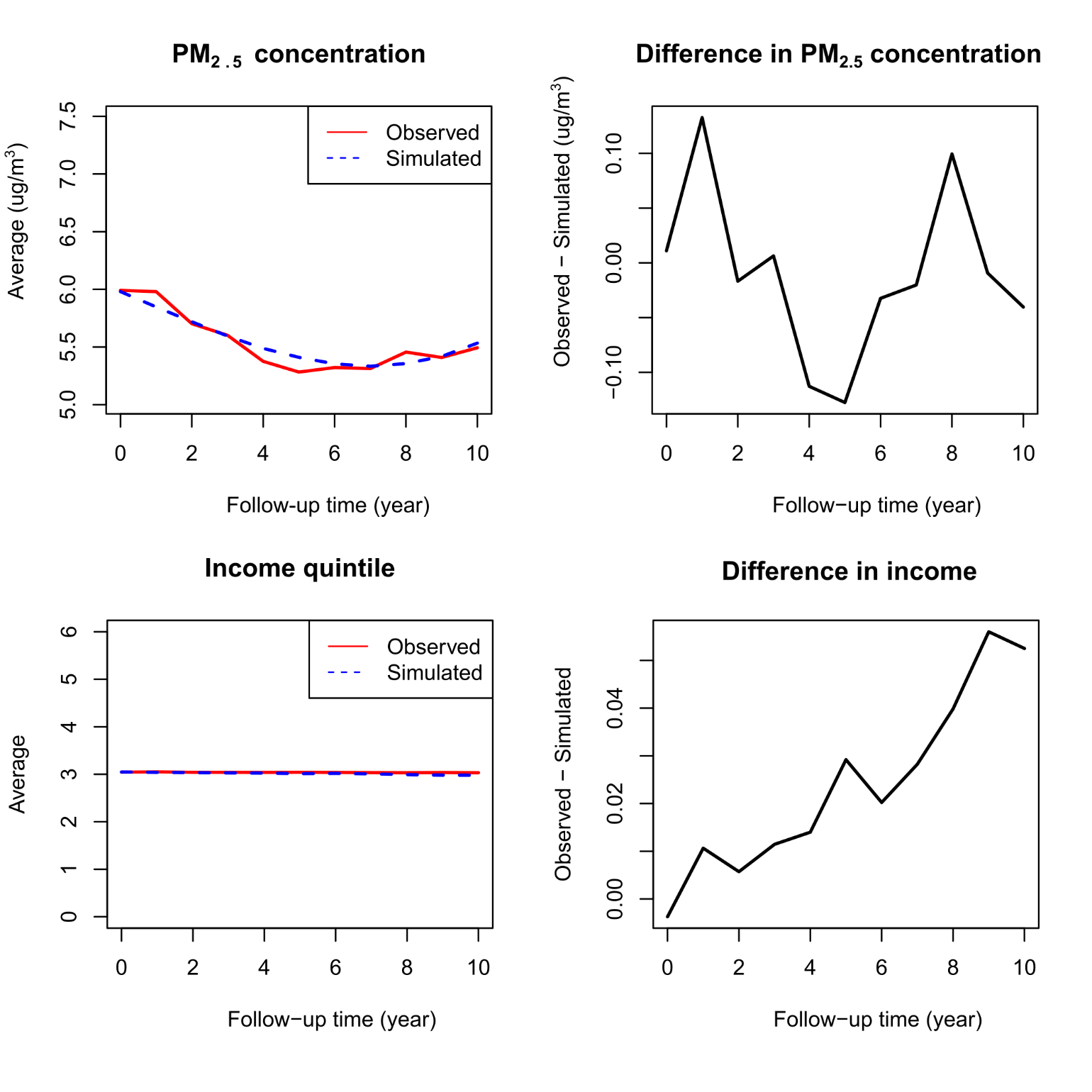

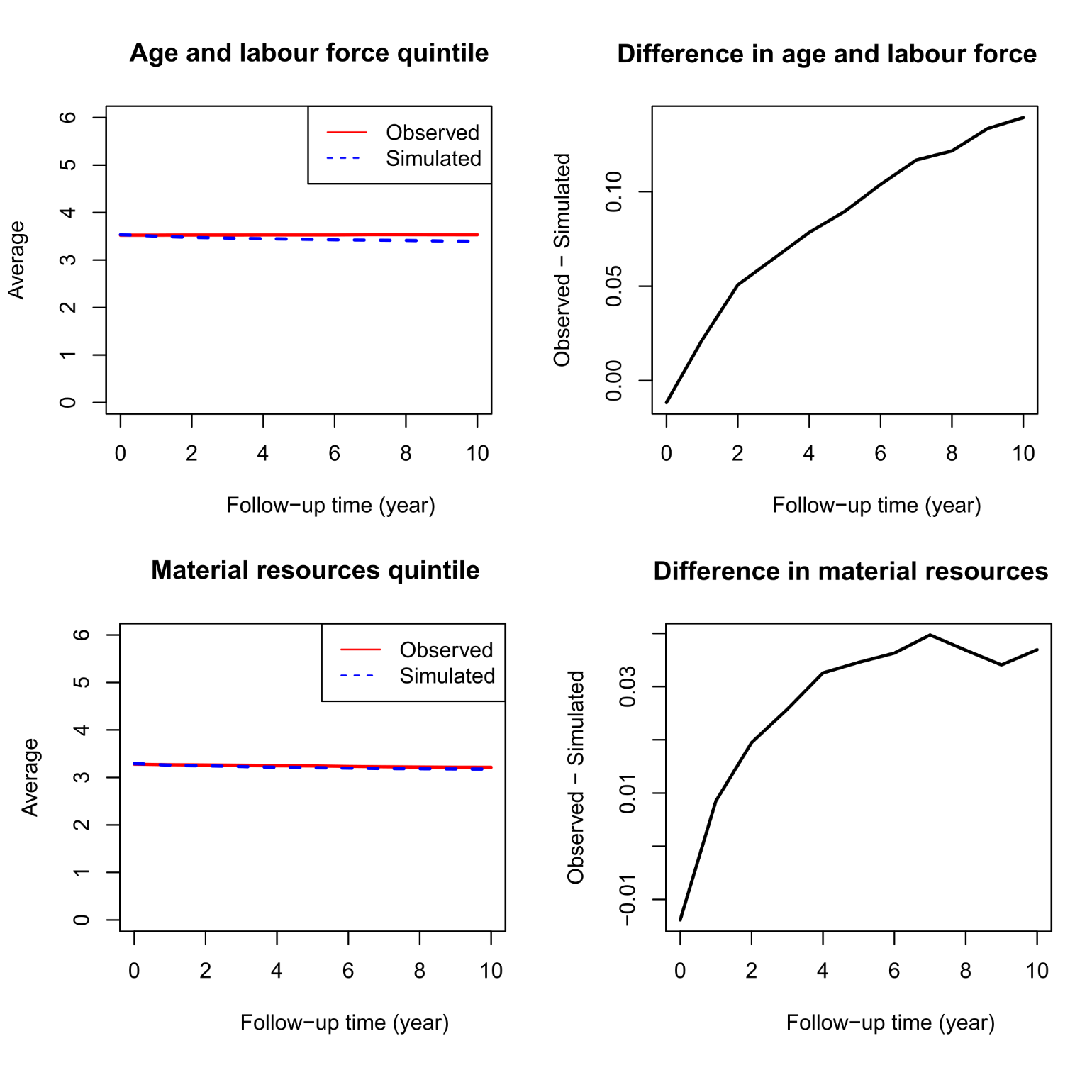

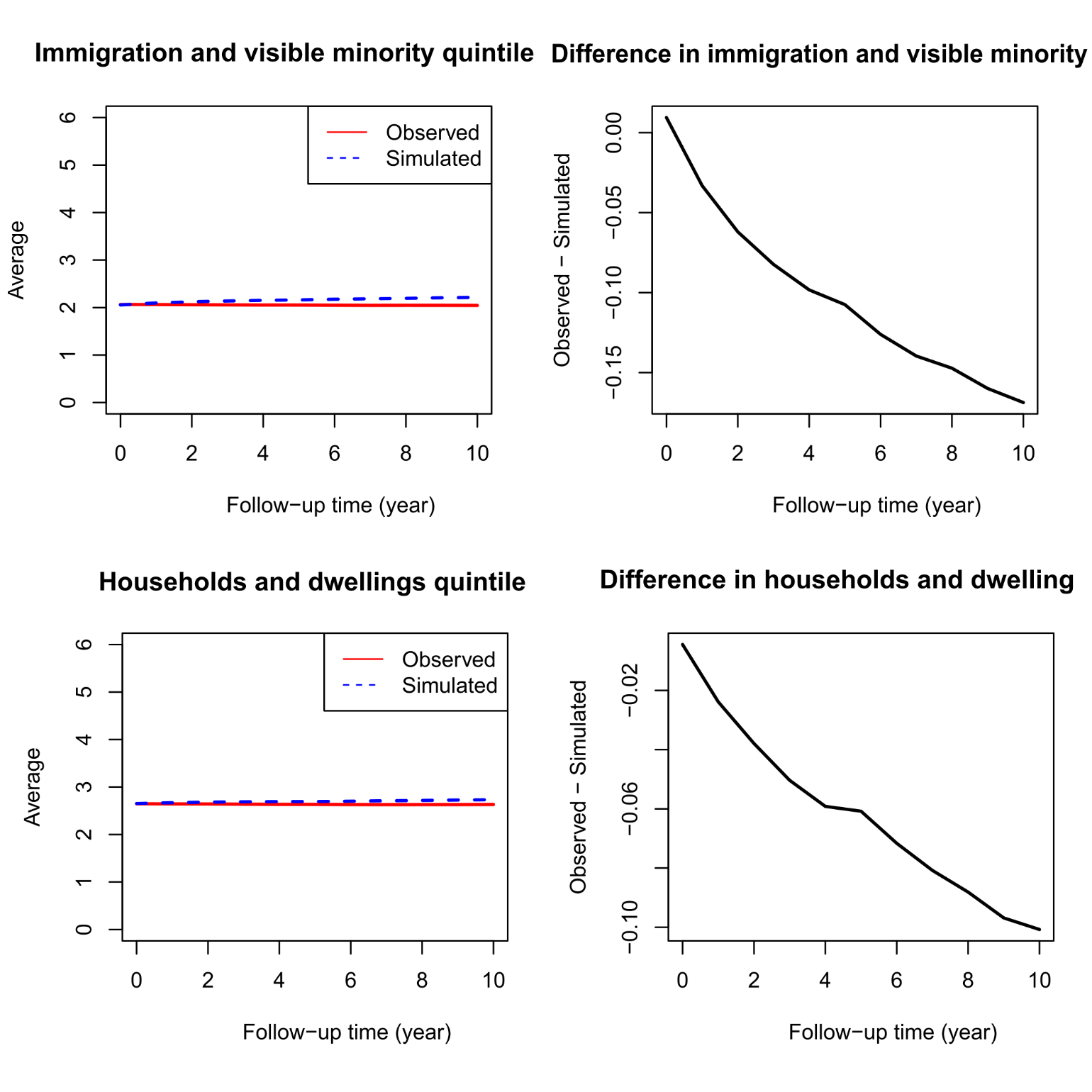

**Cycle 2003**

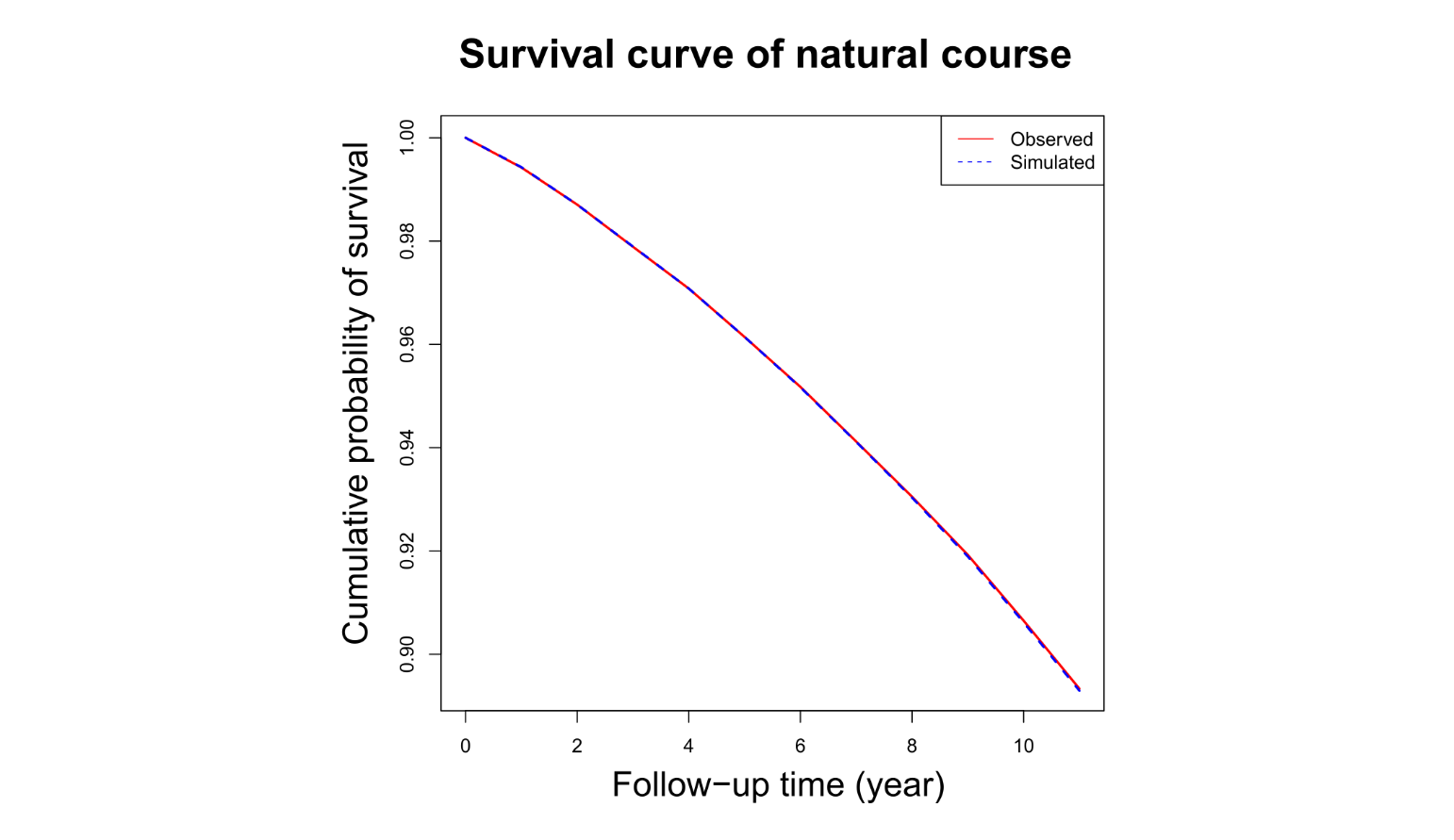

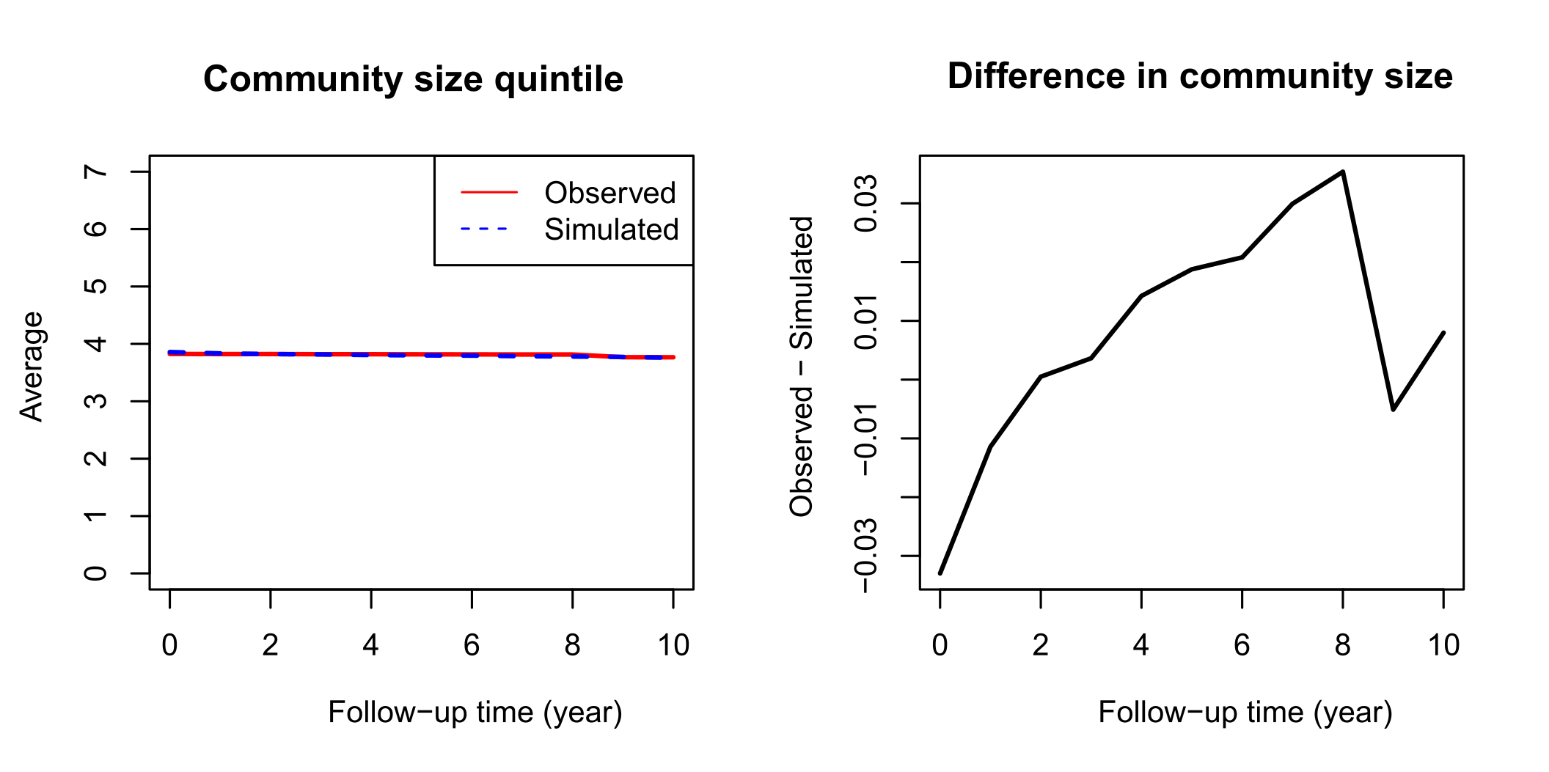

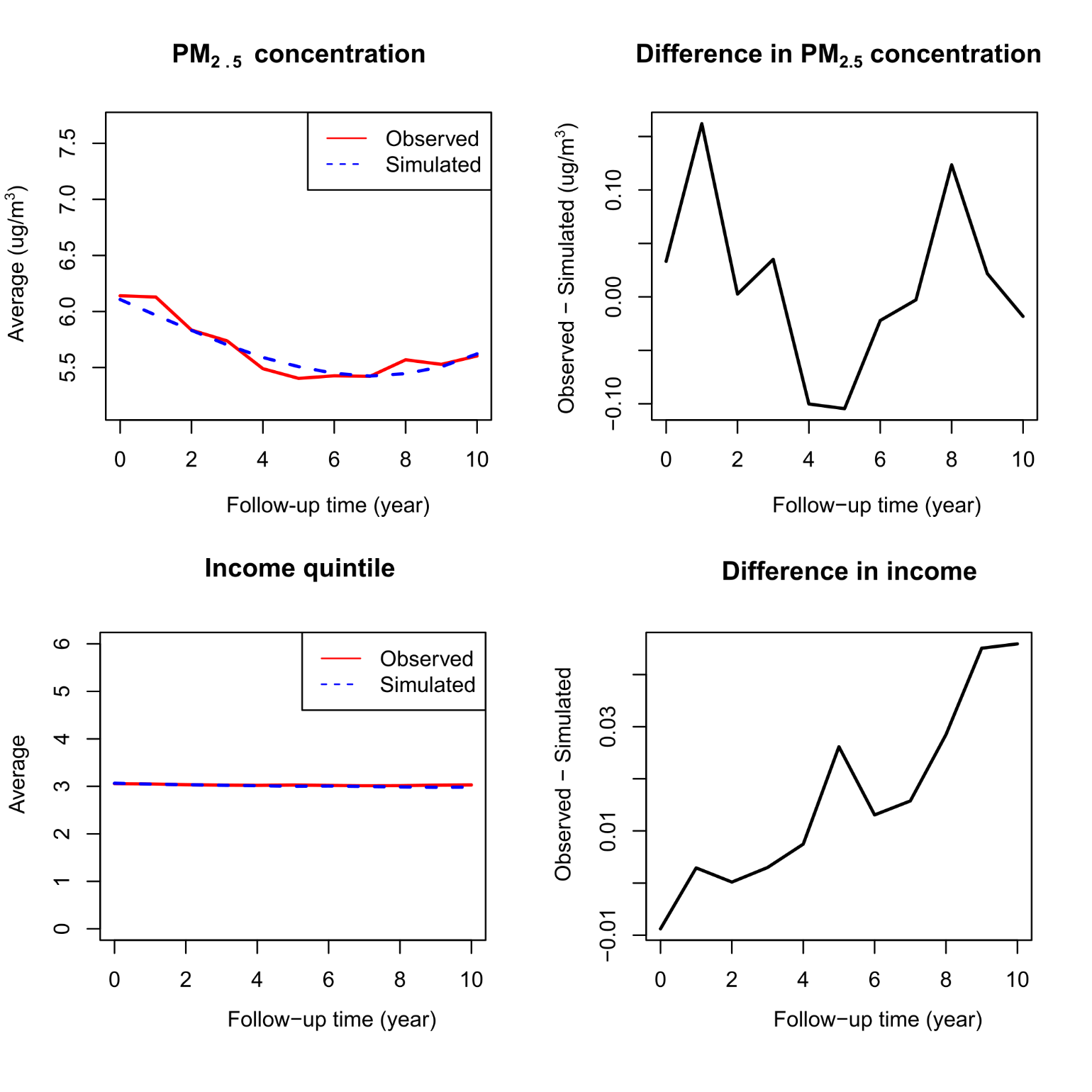

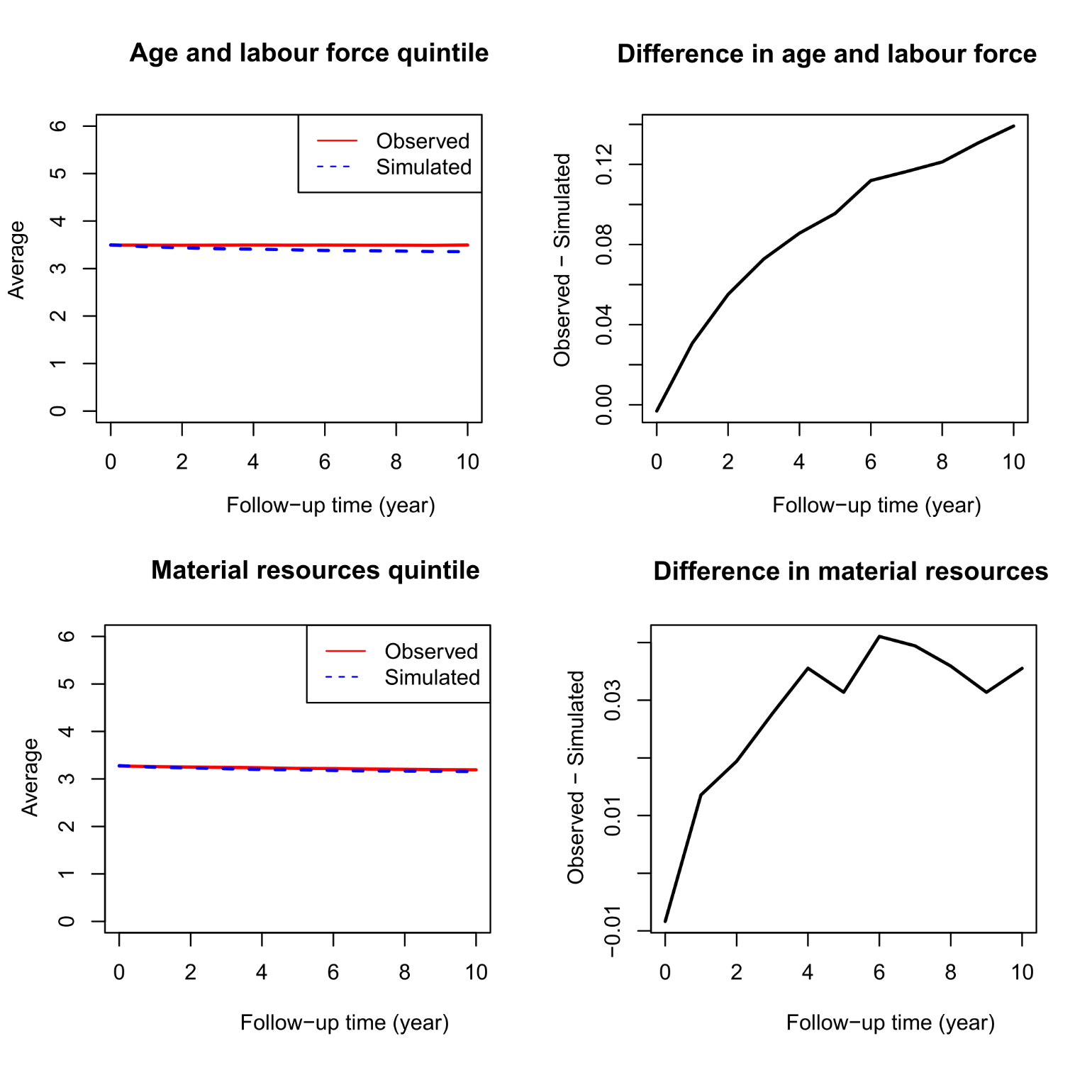

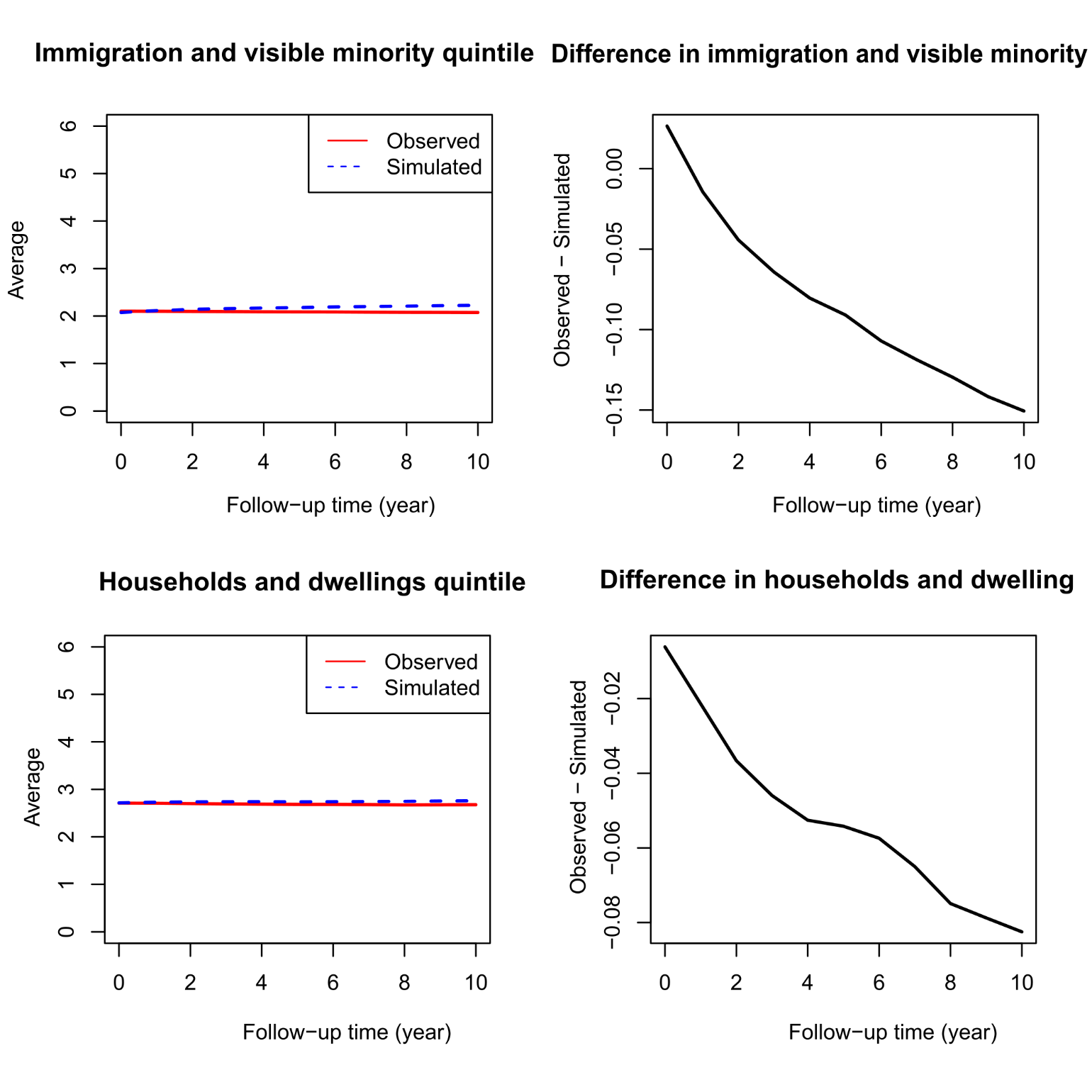

**Cycle 2005**

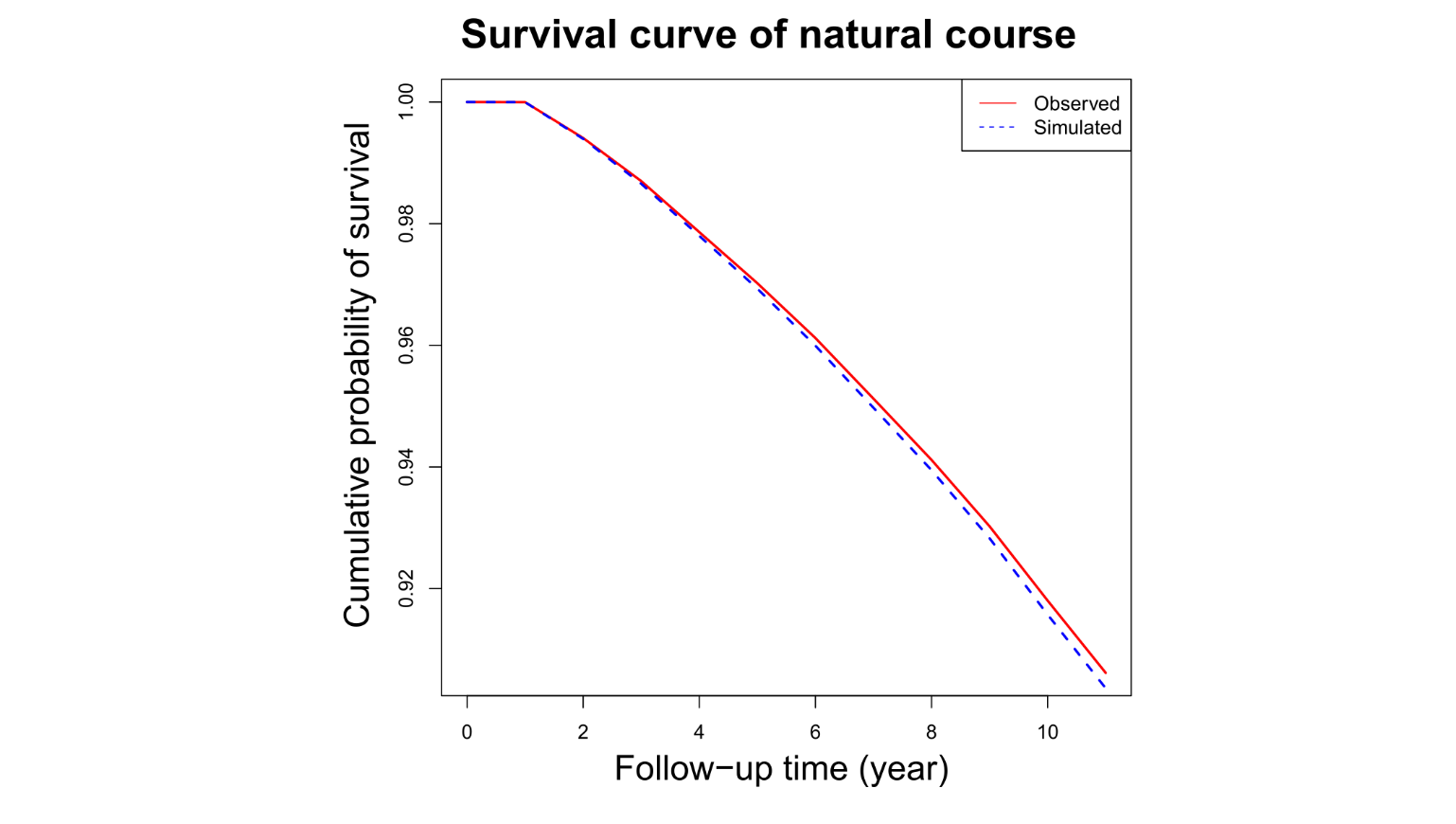

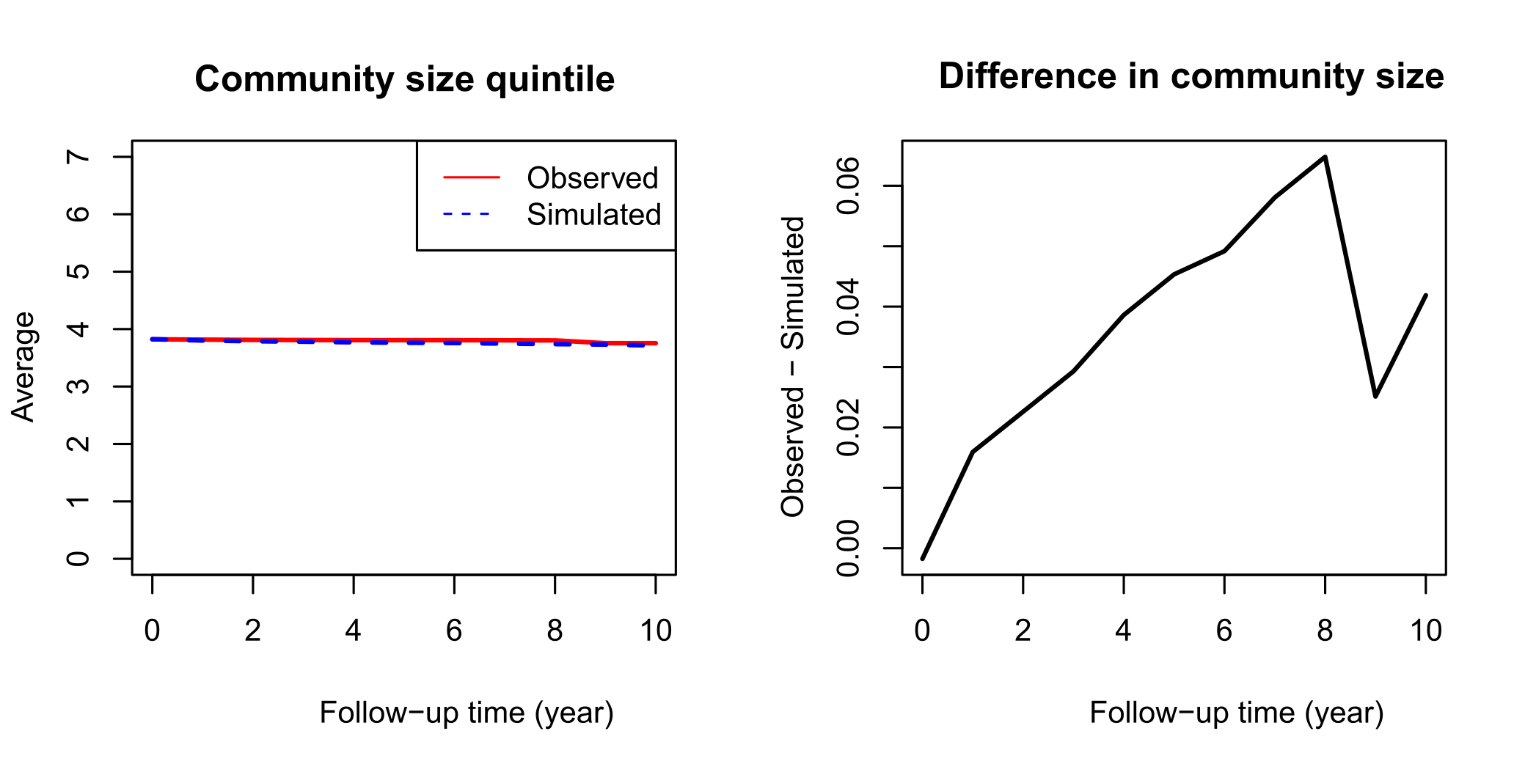

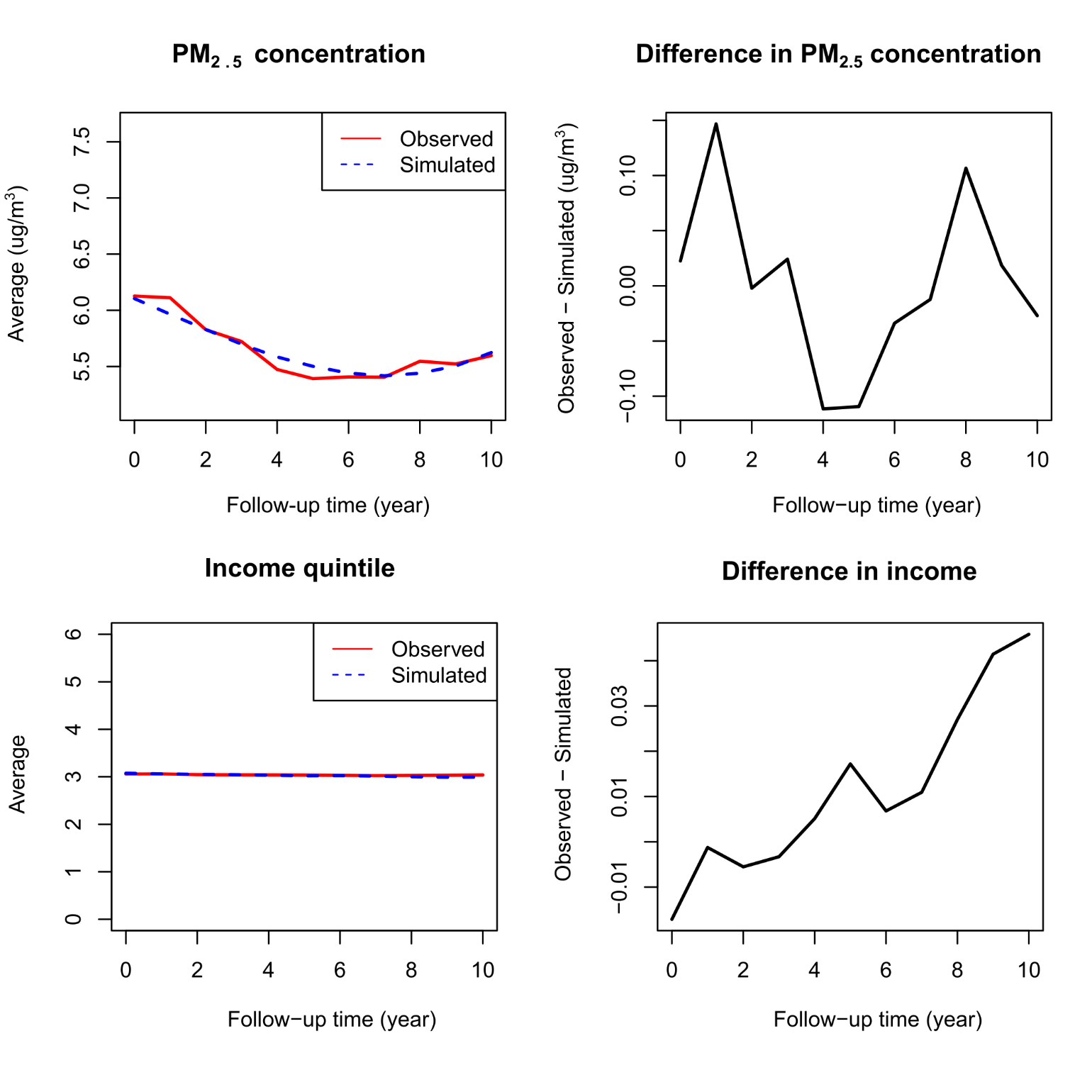

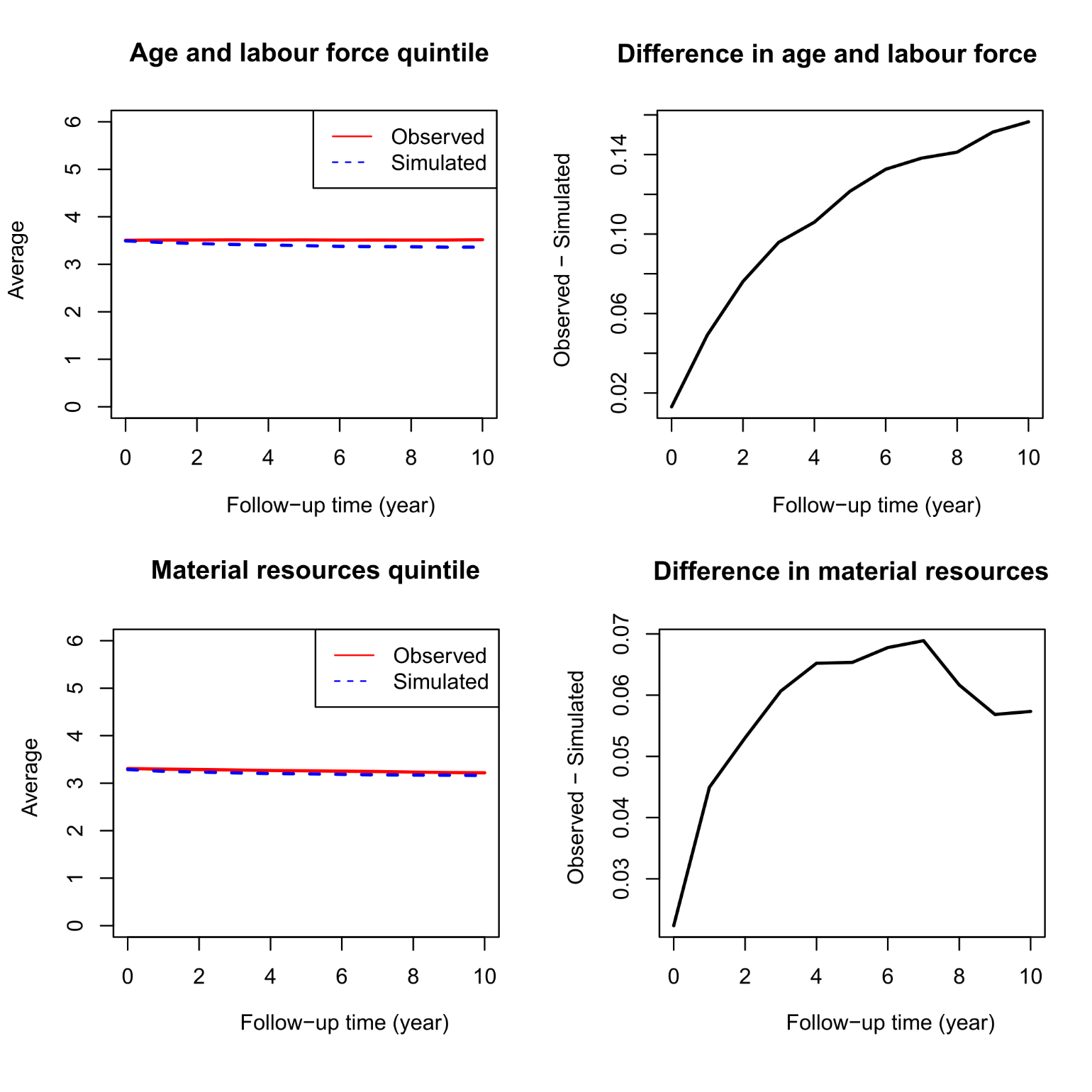

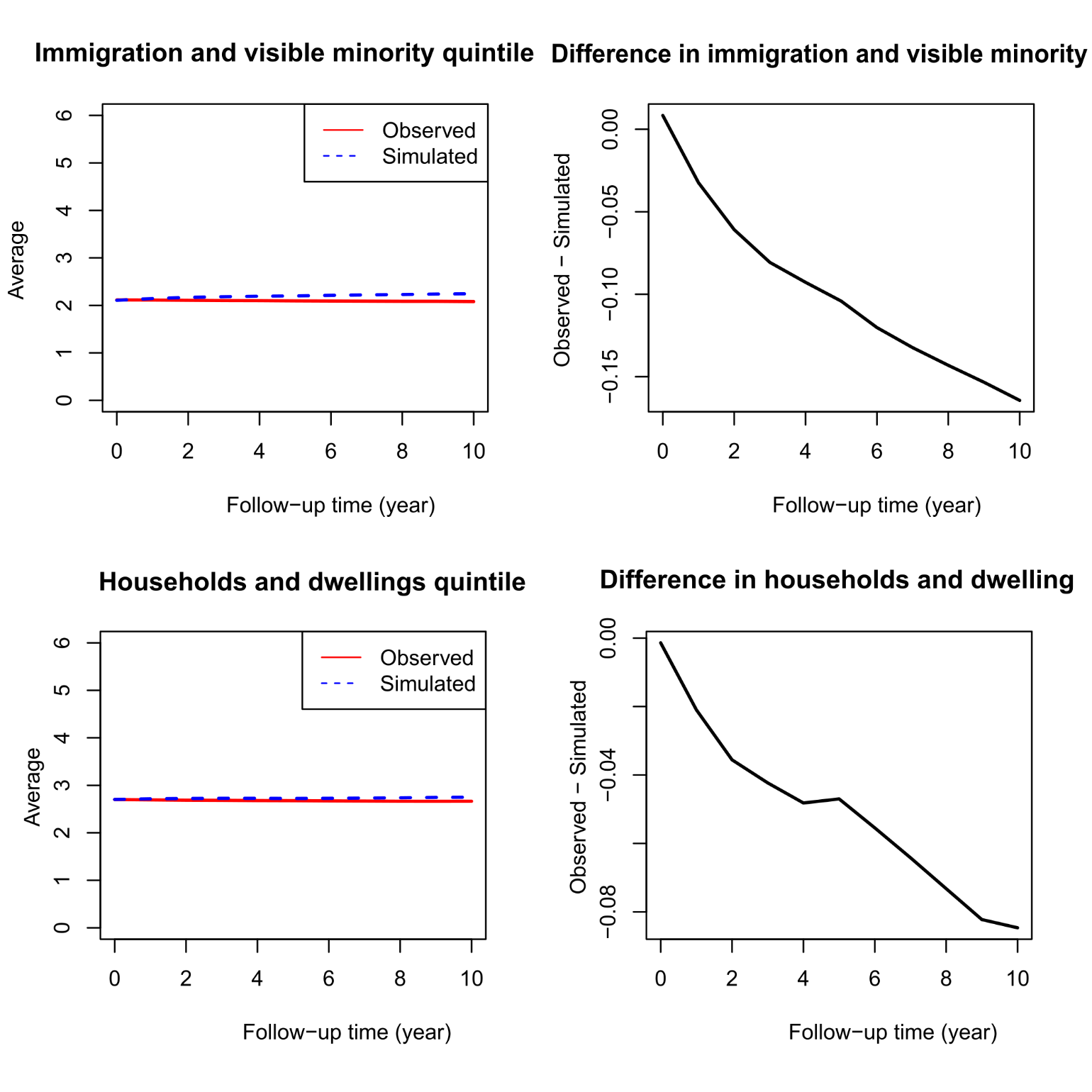
